# Monoallelic *TYROBP* deletion is a novel risk factor for Alzheimer’s disease

**DOI:** 10.1101/2024.05.09.24307099

**Authors:** Henna Martiskainen, Roosa-Maria Willman, Sami Heikkinen, Stephan A. Müller, Rosa Sinisalo, Mari Takalo, Petra Mäkinen, Teemu Kuulasmaa, Viivi Pekkala, Ana Galván del Rey, Päivi Harju, Sini-Pauliina Juopperi, Heli Jeskanen, Inka Kervinen, Kirsi Saastamoinen, FinnGen, Marja Niiranen, Sami V. Heikkinen, Mitja I. Kurki, Jarkko Marttila, Petri I. Mäkinen, Tiia Ngandu, Jenni Lehtisalo, Céline Bellenguez, Jean-Charles Lambert, Christian Haass, Juha Rinne, Juhana Hakumäki, Tuomas Rauramaa, Johanna Krüger, Hilkka Soininen, Annakaisa Haapasalo, Stefan F. Lichtenthaler, Ville Leinonen, Eino Solje, Mikko Hiltunen

**Affiliations:** Institute of Biomedicine, University of Eastern Finland; Kuopio, Finland; German Center for Neurodegenerative Diseases (DZNE); Munich, Germany; Neuroproteomics, School of Medicine and Health, Technical University of Munich; Munich, Germany; A. I. Virtanen Institute for Molecular Sciences, University of Eastern Finland; Kuopio, Finland; Neuro Center - Neurology, Kuopio University Hospital; Kuopio, Finland; Institute of Clinical Medicine - Neurology, University of Eastern Finland; Kuopio, Finland; Institute for Molecular Medicine Finland (FIMM), Helsinki Institute of Life Science (HiLIFE), University of Helsinki; Helsinki, Finland; Program in Medical and Population Genetics, Broad Institute of Harvard and MIT; Cambridge, MA, USA; Stanley Center for Psychiatric Research, Broad Institute of Harvard and MIT; Cambridge, MA, USA; Analytic and Translational Genetics Unit, Massachusetts General Hospital; Boston, MA, USA; Department of Clinical Radiology, Imaging Center, Kuopio University Hospital; Kuopio, Finland; Department of Public Health and Welfare, Finnish Institute for Health and Welfare, Helsinki, Finland; Division of Clinical Geriatrics, Center for Alzheimer Research, Department of Neurobiology, Care Sciences and Society, Karolinska Institutet, Stockholm, Sweden; Université de Lille, Inserm, CHU Lille, Institut Pasteur de Lille, LabEx DISTALZ - U1167-RID-AGE Facteurs de risque et déterminants moléculaires des maladies liées au vieillissement; Lille, France; Metabolic Biochemistry, Biomedical Centre (BMC), Faculty of Medicine, Ludwig-Maximilian University of Munich; Munich, Germany; Munich Cluster for Systems Neurology (Synergy); Munich, Germany; Turku PET Centre, University of Turku, Turku, Finland; Division of Clinical Neurosciences, Turku University Hospital, Turku, Finland; Unit of Radiology, Institute of Clinical Medicine, University of Eastern Finland; Kuopio, Finland; Department of Clinical Pathology, Kuopio University Hospital; Kuopio, Finland; Unit of Pathology, Institute of Clinical Medicine, University of Eastern Finland; Kuopio, Finland; Research Unit of Clinical Medicine, Neurology, University of Oulu; Oulu, Finland; Medical Research Center, Oulu University Hospital; Oulu, Finland; Neurocenter, Neurology, Oulu University Hospital; Oulu, Finland; Department of Neurosurgery, Kuopio University Hospital; Kuopio, Finland; Institute of Clinical Medicine, University of Eastern Finland; Kuopio, Finland

## Abstract

Biallelic loss-of-function variants in *TYROBP* and *TREM2* cause autosomal recessive presenile dementia with bone cysts known as Nasu-Hakola disease (NHD, alternatively polycystic lipomembranous osteodysplasia with sclerosing leukoencephalopathy, PLOSL). Some other *TREM2* variants contribute to the risk of Alzheimer’s disease (AD) and frontotemporal dementia, while deleterious *TYROBP* variants are globally extremely rare and their role in neurodegenerative diseases remains unclear. The population history of Finns has favored the enrichment of deleterious founder mutations, including a 5.2 kb deletion encompassing exons 1-4 of *TYROBP* and causing NHD in homozygous carriers. We used here a proxy marker to identify monoallelic *TYROBP* deletion carriers in the Finnish biobank study FinnGen combining genome and health registry data of 520,210 Finns. We show that monoallelic *TYROBP* deletion associates with an increased risk and earlier onset age of AD and dementia when compared to noncarriers. In addition, we present the first reported case of a monoallelic *TYROBP* deletion carrier with NHD-type bone cysts. Mechanistically, monoallelic *TYROBP* deletion leads to decreased levels of DAP12 protein (encoded by *TYROBP*) in myeloid cells. Using transcriptomic and proteomic analyses of human monocyte-derived microglia-like cells, we show that upon lipopolysaccharide stimulation monoallelic *TYROBP* deletion leads to the upregulation of the inflammatory response and downregulation of the unfolded protein response when compared to cells with two functional copies of *TYROBP*. Collectively, our findings indicate *TYROBP* deletion as a novel risk factor for AD and suggest specific pathways for therapeutic targeting.

**One Sentence Summary:** Nasu-Hakola disease causing *TYROBP* deletion increases the risk of Alzheimer’s disease in elderly monoallelic carriers in the Finnish population.

## INTRODUCTION

The TYRO protein tyrosine kinase-binding protein (*TYROBP*) gene encodes for DAP12 that functions as a transmembrane signaling adapter in immune cells of the myeloid lineage. Within the brain, DAP12 is expressed by microglia, where it mediates intracellular signaling from various cell surface receptors including TREM2, CR3, and SIRP1β. DAP12 has been identified as a central microglial hub in networks regulating Alzheimer’s disease (AD) pathology and microglial surveillance functions (*1, 2*).

Genetic associations with different neurodegenerative diseases highlight the indispensable role of TREM2-DAP12 signaling in brain health. Biallelic (both alleles carry a variant, homozygous or compound heterozygous) loss-of-function variants in either *TYROBP* or *TREM2* cause Nasu-Hakola disease (NHD) known also as polycystic lipomembranous osteodysplasia with sclerosing leukoencephalopathy (PLOSL). NHD is a rare recessive neurodegenerative disorder characterized by bone cysts and pathological fractures at early adulthood, followed by early onset frontotemporal type dementia and death at the middle age (*3, 4*). Additionally, biallelic *TREM2* variants contribute to recessively inherited early-onset behavioral variant frontotemporal dementia with white matter abnormalities but without bone involvement (*5–7*), while monoallelic missense variants in *TREM2* significantly increase the risk of AD (*8, 9*). Despite the close functional connection between TREM2 and DAP12, the potential contribution of *TYROBP* variants to neurodegenerative diseases remain inconclusive (*10–12*). One possible explanation for this disparity could be the extreme rarity of potentially deleterious *TYROBP* variants across most of the studied populations.

The Finnish population, characterized by a unique history of relative isolation and population bottlenecks followed by rapid expansion, harbors a relatively homogeneous genetic background with a small number of deleterious variants at higher frequencies than is commonly observed in other populations. Notably, among the deleterious founder mutations enriched in the Finnish population is a 5.2-kb deletion covering four of the five exons of *TYROBP* and causing NHD in homozygous carriers (*3, 13*). Based on the prevalence of NHD in Finland (1:500,000-1:1,000,000) (*13*) and a previous study (*11*), the frequency of monoallelic *TYROBP* deletion carriers is estimated to be around 1:300-1:500. One previous study has explored the possibility that the Finnish *TYROBP* deletion might be a risk factor for neurodegenerative diseases in the monoallelic carriers but found no association (*11*). However, the result may be considered inconclusive due to the limited number of identified *TYROBP* deletion carriers (*11*). Studies in larger cohorts have been hindered by the difficulty to detect large structural variants in generally used genotyping array data, which mostly comprises single nucleotide variants (SNVs).

We hypothesized that assessing *TYROBP* deletion in FinnGen, a large, well-characterized biobank-scale cohort representing 10 % of the Finnish population (*14*), would provide a more comprehensive understanding on the potential phenotype effect of the monoallelic *TYROBP* deletion and, more broadly, on the effect of partial *TYROBP* loss. The feasibility of FinnGen for such analyses was recently demonstrated by a study that identified novel significant phenotype associations in the monoallelic state for many known recessive disease-causing variants (*15*).

In the present study, we identified two SNVs as genetic proxies for the Finnish *TYROBP* deletion and confirmed that this deletion is a Finnish founder mutation. Importantly, our findings revealed that the Finnish *TYROBP* deletion associates with and increased risk and earlier onset of dementia and AD in the monoallelic carriers. Furthermore, we present a case of monoallelic *TYROBP* deletion carrier exhibiting cystic bone lesions reminiscent of those detected in NHD patients, but without cognitive symptoms. Finally, we demonstrated that *TYROBP* deletion in mono- and biallelic carriers induces functional alterations in microglia-like cells *in vitro*.

## RESULTS

### Intronic SNVs in *TYROBP* and *NFKBID* are proxy markers for the Finnish *TYROBP* deletion

To identify genetic proxy markers for the Finnish *TYROBP* deletion, we performed whole genome sequencing (WGS) analysis on three NHD patients unrelated up to at least 3^rd^ degree. WGS confirmed biallelic deletion spanning approx. 5.2 kb and encompassing the exons 1-4 of *TYROBP* in all three NHD patients (Fig. 1A).

**Fig. 1.**
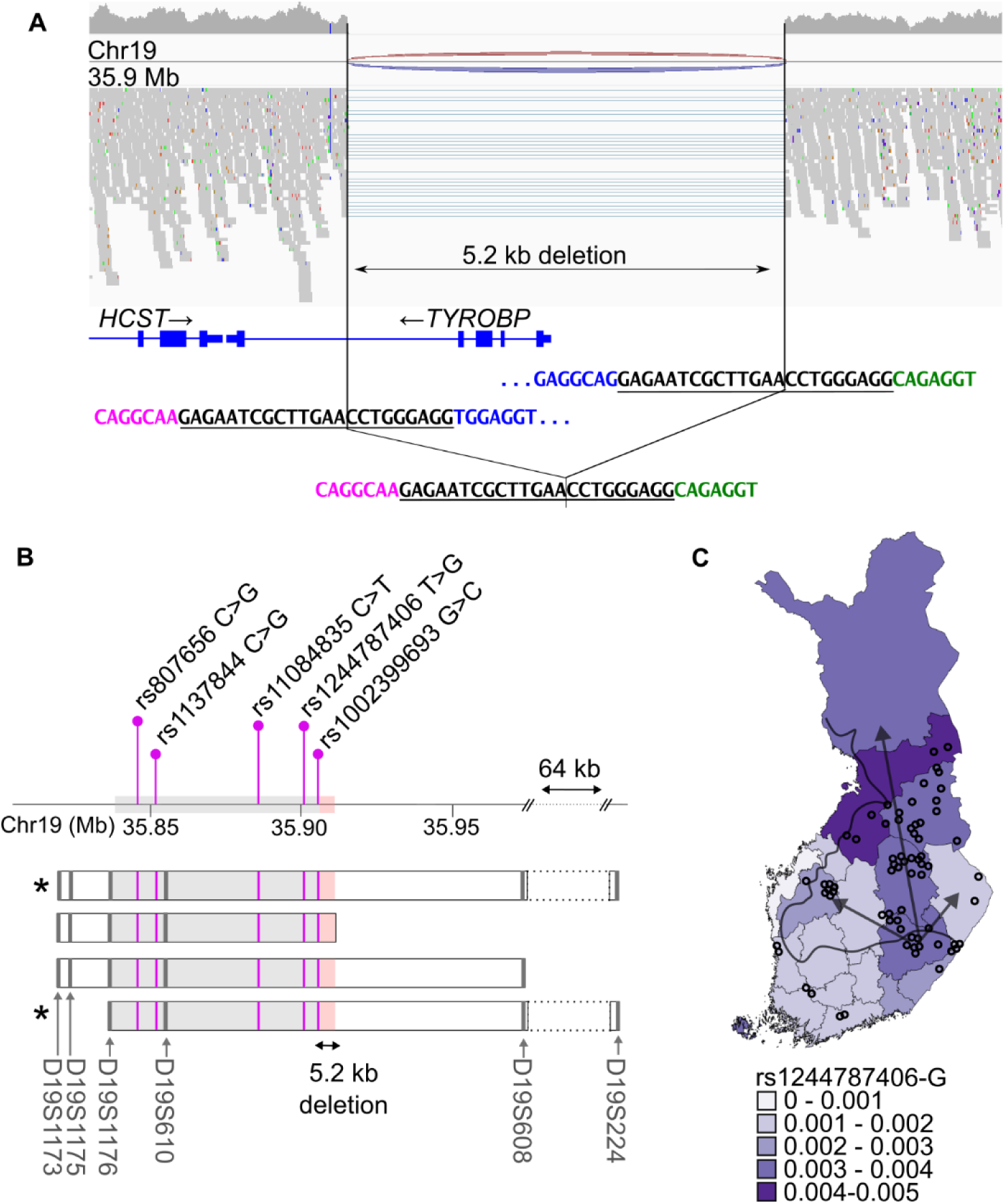
The Finnish 5.2 kb *TYROBP* deletion. **(A)** WGS data show homozygous 5.2-kb deletion encompassing *TYROBP* exons 1-4 in a Finnish NHD patient. The horizontal lines indicate WGS reads spanning across the deletion. The deletion breakpoints are located within a 23-bp identical sequence (black, underlined). **(B)** Schematic of the *TYROBP* deletion founder haplotype. The boxes indicate haplotype blocks identified using microsatellite markers (grey bars) (13). The grey shading indicates the haplotype region shared by all Finnish NHD patients, while the red shading indicates the 5.2-kb *TYROBP* deletion. Asterisks denote the haplotypes detected in the NHD patients in the current study. Purple lines indicate the SNVs identified in the current study within the shared haplotype region. **(C)** Regional allele frequency of rs1244787406-G in Finland. The black line separates the early settlement region along the southern and western coastlines from the late settlement region that was permanently inhabited from the 16th century onwards by internal migration (arrows) mainly from the current South Savo region in the southeastern Finland. The circles represent the birth places of the grandparents of the Finnish NHD patients according to (17). The highest present day allele frequencies of the *TYROBP* deletion proxy marker rs1244787406-G are detected in the late settlement area and coincide with the previously reported regional enrichment.

The haplotype containing the Finnish *TYROBP* deletion shared by all Finnish NHD patients has been shown to cover at least 68.915 kb immediately upstream of the deletion (Fig. 1B) (*3, 13*). We focused our search of *TYROBP* deletion-associated single nucleotide variants (SNVs) to this shared haplotype region, even though the shared area of homozygosity in the NHD patients included in our study was considerably larger, at least 1.95 Mb, spanning both up- and downstream of the deletion. A total of seven SNVs homozygous for the minor allele in the NHD patients were detected in the shared haplotype region (Table S1). All the identified variants were intronic or intergenic with minor allele frequency (MAF) of 0.0031-0.79 in the Finnish population (*16*). Interestingly, the two variants closest to the deletion, rs1002399693 (19:35,905,673 G>C in *TYROBP* intron 4, MAF=0.0032) and rs1244787406 (19:35,901,079 T>G in *NFKBID* intron 1, MAF=0.0031) were enriched in the Finnish population compared to non-Finnish Europeans, and had MAF matching the estimated carrier frequency of the Finnish 5.2-kb *TYROBP* deletion (*11*).

Next, we explored the *TYROBP* deletion haplotype and individual SNVs in the FinnGen data. Two variants were not available in FinnGen, and thus, haplotype based on imputed, phased genotypes of five SNVs was used to identify putative *TYROBP* deletion carriers. Among the 520,210 individuals, the haplotype consisting of five SNVs identified 2,231 putative *TYROBP* deletion carriers, corresponding to MAF= 0.0021. All the same 2,231 individuals and one additional individual were identified when using only the SNV rs1244787406 T>G or rs1002399693 G>C, suggesting that either of these SNVs alone could be used as a proxy marker for the 5.2-kb *TYROBP* deletion. Since the imputation Info score for these variants in FinnGen is only 0.84, we confirmed the presence of both variants in the WGS data from three Finnish NHD patients and one monoallelic carrier of the 5.2-kb *TYROBP* deletion, and the rs1244787406 T>G by Sanger sequencing in three NHD patients and four monoallelic carriers (Fig. S1A-B).

Furthermore, we confirmed the presence of monoallelic *TYROBP* deletion in 49 FinnGen subjects with imputed rs1244787406-G and rs1002399693-C alleles with high (>0.99) genotype probability. One sample with low genotype probability (0.52) was found to be homozygous for *TYROBP* common variant (Fig. S1C). The rs1244787406 T>G was used as the proxy marker for *TYROBP* deletion in all the further analyses conducted in FinnGen data.

We next calculated the frequency by the region of birth for the identified putative *TYROBP* deletion carriers using MAFs for rs1244787406-G in FinnGen. There was no statistically significant difference in the regional distribution (chi-square test, n.s.). The highest frequencies were observed in North Ostrobothnia (MAF=0.0040) and North Savo (MAF=0.0036), while the lowest were found in Ostrobothnia (MAF=0.00063) (Fig. 1C). This regional enrichment is consistent with the previously reported birthplaces of Finnish NHD patients, their parents and grandparents (*13, 17*). Together, these results suggest that the rs1244787406-G can be reliably used as a proxy marker to identify carriers of the Finnish founder mutation, the 5.2-kb *TYROBP* deletion.

### Monoallelic *TYROBP* deletion associates with an increased risk and earlier onset-age of dementia and AD

To elucidate the phenotypic effect of the monoallelic *TYROBP* deletion, we performed phenome-wide association scan (PheWAS) against 2489 clinical core endpoints available in FinnGen. A complete list of endpoints and their definitions can be found at https://www.finngen.fi/en/researchers/clinical-endpoints. PheWAS for rs1244787406-G indicated increased risk of five endpoints related to dementia and Alzheimer’s disease (Fig. 2A, Table S2).

**Fig. 2.**
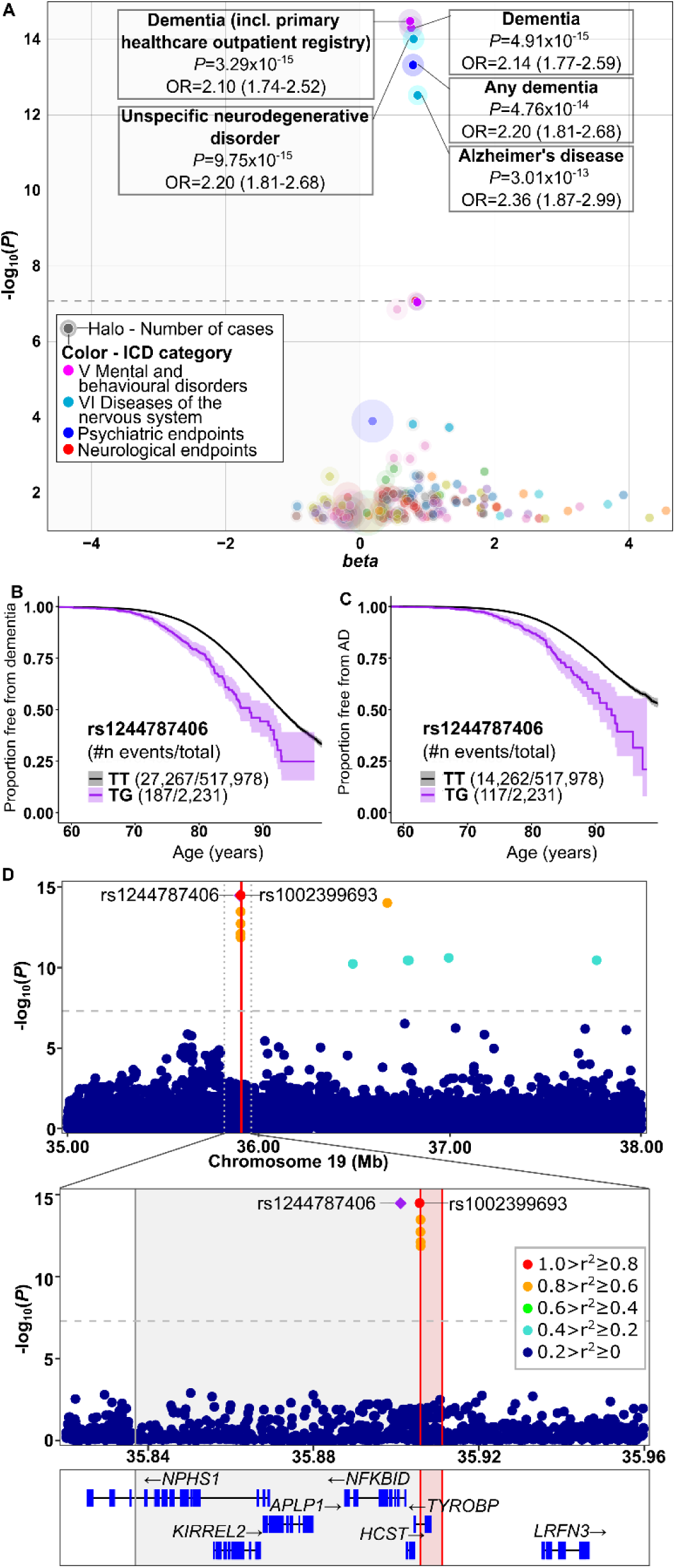
Phenotype associations for the *TYROBP* deletion proxy marker rs1244787406-G and regional association plot of the *TYROBP* locus. **(A)** Phenotype association study covering 2489 endpoints indicates significantly increased risk for dementia and AD among carriers of rs1244787406 minor G allele. The dashed line represents genome-wide significance threshold *P*=5×10^−8^. **(B-C)** Kaplan-Meier survival plots showing the proportion free from dementia (B) and AD (C) among rs1244787406-G carriers (purple line) and non-carriers (black line). Shading indicates 95 % confidence intervals. X-axis indicates age at the first diagnosis for cases and age at the end of follow-up for controls. **(D)** Regional association plot of the *TYROBP* locus shows the negative log10-transformed P-values on the y axis for the endpoint ‘Dementia (including primary healthcare outpatient registry)’ derived from FinnGen. The vertical dashed line represents genome-wide significance threshold *P*=5×10^−8^. Each dot represents an individual SNV, and the dot color represents LD with the LD reference variant (purple diamond). The dotted line in the upper panel delineates the area shown in the lower panel. The red vertical lines indicate the 5.2 kb *TYROBP* deletion break points while the shaded grey area represents the *TYROBP* deletion associated shared haplotype region in the lower panel.

To investigate whether the rs1244787406 T>G affects the age of disease onset, we generated Kaplan-Meier survival curves for dementia in general (including AD) (Fig. 2B) and more specifically for AD only (Fig. 2C) illustrating disease free survival from birth to the age at first diagnosis for cases and the age at the end of follow-up for controls. For both dementia and AD, the *TYROBP* deletion proxy marker rs1244787406-G significantly lowered the age at first diagnosis (log-rank test, Chi-squared=86.2, df=1, P<2×10^−16^; Fig. 2B-C). Median age for onset of dementia was 76.5 (interquartile range 72.1-81.9) years for rs1244787406-G carriers and 79.0 (73.5-83.7) years for non-carriers; and for AD 78.61 (interquartile range 73.8-83.3) years for rs1244787406-G carriers and 80.5 (75.3-85.0) years for non-carriers.

To determine whether the association with dementia and AD was driven by the *TYROBP* deletion proxy markers or other variants in the locus, we produced regional association plots of the *TYROBP* locus (Fig. 2D). The two identified *TYROBP* deletion proxy markers were in high linkage disequilibrium (LD; r^2^=1), had the highest association signal in the locus and were the only significantly associated variants within the known shared *TYROBP* deletion haplotype. Other variants within the *TYROBP* locus were in weaker LD (r^2^<0.8) to the lead variants and were located within the deleted sequence or in the haplotype region shared only by some, but not all *TYROBP* deletion carriers (Fig. 2D, Fig. 1A).

To better understand the role of the variants located within the deleted region detected in FinnGen, we further analysed our WGS data from three NHD patients and one monoallelic deletion carrier. Variants in the region 19:35905884-19:35905910 were found to be artefacts arising from the *TYROBP* deletion (Fig. S2). The deletion breakpoints are located within 120 bp repeated sequence that is almost identical on both sides of the deletion but differs on 15 bases along the repeat. Due to the repeated sequence, reads spanning the deleted area are mapped to one side of the deletion only and the different bases appear as SNVs. However, when large deletions are allowed in the alignment, the reads are aligned correctly and span the deleted sequence, indicating that these SNVs were artefacts arising from the deletion (Fig. S2).

Since the *TYROBP* gene resides 9 Mb upstream of *APOE* in chromosome 19, we sought to determine whether the *APOE* ε4 allele, the strongest known genetic risk factor for AD, was co-inherited with *TYROBP* deletion. To this end, we derived *TYROBP* deletion proxy marker and *APOE* ε4 (rs429358) genotypes from the FinnGen genotype data. *TYROBP* deletion carriers were less likely to have the *APOE* ε4 allele as compared to *TYROBP* deletion non-carriers (30.2 % vs 32.8 %, Chi-squared=7.152, df=1, P=0.0075). These results suggest that *TYROBP*-deletion is not co-inherited with *APOE* ε4 and thus increases the risk of AD and dementia independently of *APOE* ε4.

### Monoallelic *TYROBP* deletion induces cystic bone lesions

Bone cysts in the wrists and ankles are an integral feature of NHD caused by the loss of *TYROBP* or *TREM2*. One report has described osteolytic lesions in monoallelic siblings of an NHD patient carrying two different *TYROBP* missense variants (Shboul et al., 2019). We report here a case of a monoallelic 5.2-kb *TYROBP* deletion carrier with classic painful NHD bone cysts but no detectable brain pathology, cognitive findings, or neuropsychiatric symptoms in their third decade of life. Monoallelic 5.2-kb *TYROBP* deletion carrier status was detected in a clinical genetic analysis and confirmed with deletion-specific PCR (Fig. S1C). To exclude the possibility of NHD caused by compound heterozygosity, i.e. biallelic state due to the presence of two different NHD-causing variants, WGS analysis was carried out. Only common variants (MAF 0.7-0.71 in the Finnish population according to gnomAD) in *TYROBP* and no variants in *TREM2* were identified in the WGS, confirming that the only NHD-associated variant in the patient was monoallelic 5.2-kb *TYROBP* deletion.

The bone and joint structures of the monoallelic *TYROBP* deletion carrier were otherwise unremarkable, except for two small cystic-appearing translucencies in the wrists (lunates) and slight ulnar minus variance (Fig. 3A), and cystic-appearing translucencies in the left calcaneus and the left distal tibia (Fig. 3B), bearing resemblance to the classical findings in NHD. The carrier’s sibling with biallelic *TYROBP* deletion and diagnosed with NHD showed multiple, widespread cystic bone lesions in the hands, wrists, ankles, and feet, with pathological fractures in the right wrist, all consistent with NHD (Fig. 3C-D). No structural or signal pathology was observed on MRI (Fig. 3E) and the brain structural volumes showed no quantitative evidence for atrophic brain pathology in the monoallelic *TYROBP* deletion carrier. The sibling with NHD, however, showed clear findings consistent with NHD: bilateral calcifications of the basal ganglia and frontal white matter, and marked cortical and temporomesial atrophy with rapid progression over time (Fig. 3F). These findings indicate that monoallelic *TYROBP* deletion can cause cystic bone lesions similar to those seen in NHD patients.

**Fig. 3.**
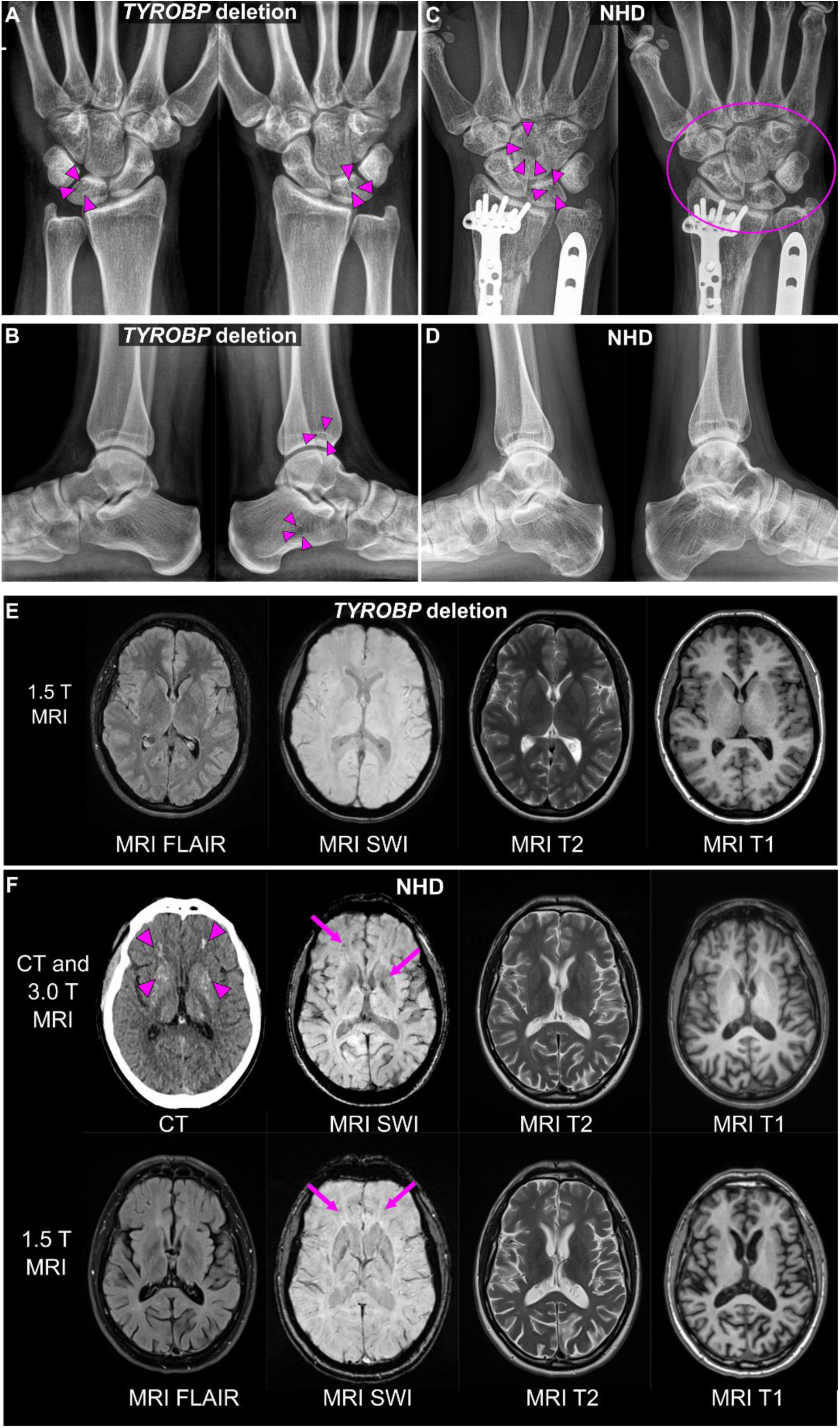
Clinical imaging of a monoallelic *TYROBP* deletion carrier and an NHD patient. **(A-B)** Radiographs of wrists and ankles of the monoallelic *TYROBP* deletion carrier in their thirties show small cystic-appearing translucencies in the lunate bones of the wrists and the left calcaneus and distal tibia of the ankle, as delineated by arrowheads **(C)** Post-traumatic and postoperative radiographs of the NHD patient’s right wrist over time span of 5 years. The bone structure is patchy already in the earlier radiograph (on the left), with cystic-appearing translucencies notably in the lunate and capitate. Five years later the primary findings are even more pronounced and all carpal bones as well as the distal radius and ulna present as abnormal (within the ellipse). **(D)** Radiograph of the NHD patient’s ankles shows numerous abnormal translucencies in the distal heads of the tibia and fibula, talar, calcaneal, and cuboid bones. **(E)** Magnetic resonance images (MRI) of the monoallelic *TYROBP* deletion carrier. No signal or structural pathologies are observable, and there is no evidence of marked atrophy on any of the contrast sets (FLAIR, T1- or T2-weighted, or susceptibility weighted images (SWI)). **(F)** Computed tomography (CT) and MRI images of the brain of the NHD (1.5 T MRI on the lower row three years after the initial CT and 3.0 T scan). CT scan shows bilateral calcifications of the basal ganglia and frontal white matter as hyperdensities (purple arrowheads). Some punctate signal voids can be observed in the corresponding regions on MRI by SWI both at 3.0T and at 1.5T field strengths (arrows). FLAIR, T2- or T1-weighted images show no obvious signal pathology. Marked cortical and temporomesial atrophy is however present and shows rapid progression over time.

### Monoallelic *TYROBP* deletion may affect β-amyloid pathology in the frontal cortex and cerebrospinal fluid (CSF)

AD-associated variants in *TREM2*, encoding the receptor associated with *TYROBP*, are known to reduce the clustering of microglia around the Aβ plaques (*18*). To examine whether the *TYROBP* loss phenocopies the AD-associated *TREM2* variants, we utilized immunohistochemical staining in frontal cortex biopsies to assess microglial clustering around the Aβ plaques (Fig. 4A) in one monoallelic *TYROBP* deletion carrier and ten non-carriers with different *APOE* genotypes from Kuopio normal pressure hydrocephalus (NPH) registry, a cohort containing surgical frontal cortex biopsies collected during ventriculoperitoneal shunt placement to treat suspected NPH (*19*). In addition, AD-related CSF biomarkers Aβ42, Tau phosphorylated at Serine 181 (p-Tau181) and total Tau were assessed in the same individuals. There were no major differences in microglial clustering or CSF biomarkers between the *TYROBP* deletion carrier and the non-carriers. The monoallelic *TYROBP* deletion carrier had more plaques per mm^2^ of tissue as compared to the non-carriers (Fig. 4B), but Aβ plaque size and microglial clustering around the Aβ plaques was within the same range as in the non-carrier individuals (Fig. 4C-E). Analysis of AD related biomarkers in the CSF indicates significantly decreased Aβ42 levels among the *APOE* ε3ε4 carriers compared to *APOE* ε3ε3 carriers (Fig. 4F) but no differences in the levels of p-Tau181 or total Tau between the genotypes (Fig. 4G-H). The CSF biomarker concentrations for the monoallelic *TYROBP* deletion carrier were within the same range as for the other *APOE* ε3ε4 carriers. Compared to the *APOE* ε3ε3 individuals, p-Tau181/Aβ42 ratio showed a light, but not statistically significant increase in the *APOE* ε3ε4 and *TYROBP* deletion/*APOE* ε3ε4 carriers (Fig. 4I), while there were no differences between the genotypes in the p-Tau181/total Tau ratio (Fig. 4J). An increased number of Aβ plaques was observed in one monoallelic *TYROBP* deletion carrier, but no differences in the number of plaque-associated microglia or AD-related CSF biomarkers were detected when compared to controls, suggesting that the monoallelic *TYROBP* deletion may not have major effects on AD-related neuropathology.

**Fig. 4.**
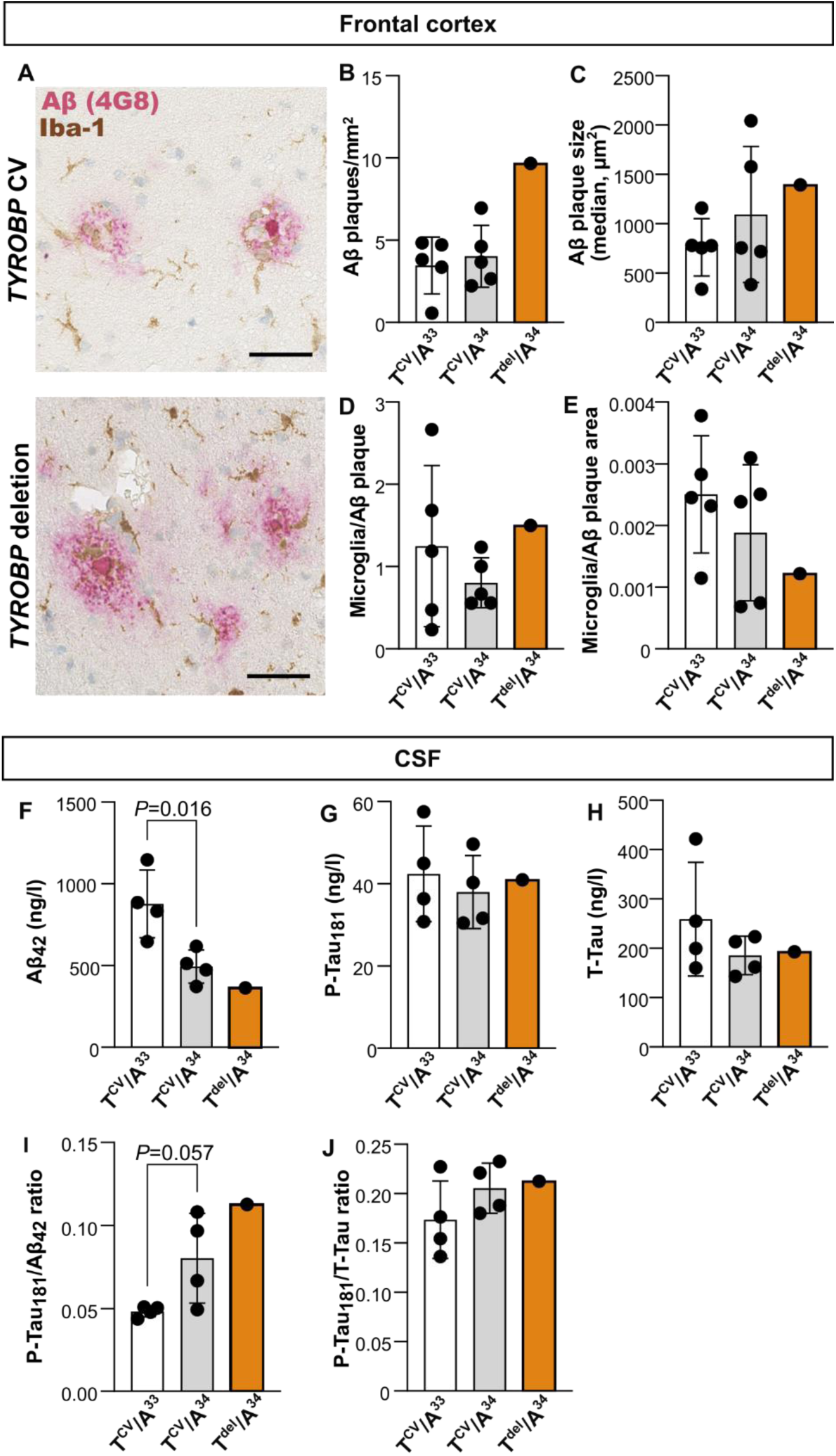
Effect of the Finnish 5.2-kb *TYROBP* deletion on Alzheimer’s disease-related β-amyloid and Tau pathology in the frontal cortex and CSF. **(A)** A representative image of frontal cortex biopsy immunostained for microglia (Iba-1, brown) and Aβ (red). Scale bar 50 µm. **(B-E)** Quantifications of immunohistochemical images shown in (A); n=1-5 individuals per genotype. **(F-J)** Quantification of Aβ and Tau biomarkers in the CSF; n=1-4 individuals per genotype. Data are shown as mean ± SD. Independent samples T-test. T^CV^, *TYROBP* common variant; T^del^, monoallelic *TYROBP* deletion; A^33^, *APOE* ε3ε3; A^34^, *APOE* ε3ε4.

### *TYROBP* deficient monocytes can be differentiated into monocyte-derived microglia-like cells (MDMi)

To understand the effects of the *TYROBP* deletion in myeloid lineage cells, we obtained monocytes from NHD patients with biallelic *TYROBP* deletion (hereafter referred to as NHD), monoallelic *TYROBP* deletion carriers, and control individuals (Fig. 5A). Since microglia are the primary myeloid cell type in the brain and the only CNS resident cell type expressing DAP12 protein derived from *TYROBP*, we next differentiated the monocytes into monocyte-derived microglia-like cells (MDMi) by culturing the cells for 11-13 days in the presence of M-CSF, GM-CSF, NGF-β, CCL2, and IL-34, similarly to previous publications (*20, 21*). First, we confirmed that the DAP12 protein was robustly expressed in the monocytes and MDMi, and that both the monocytes and MDMi derived from the *TYROBP* deletion carriers displayed reduced DAP12 levels when compared to controls (Fig. 5B). A similar effect was observed at RNA level in both monocytes and MDMi, where the monoallelic *TYROBP* deletion carriers had reduced TYROBP RNA levels as compared to the control individuals carrying the common variant of *TYROBP*, while no TYROBP RNA was detected in cells derived from the NHD patients homozygous for the *TYROBP* deletion (Fig. 5C). These results suggest a gene dose-dependent reduction in *TYROBP*/DAP12 levels in myeloid cells derived from individuals with one or two copies of *TYROBP* deletion compared to individuals with *TYROBP* common variant.

**Figure 5.**
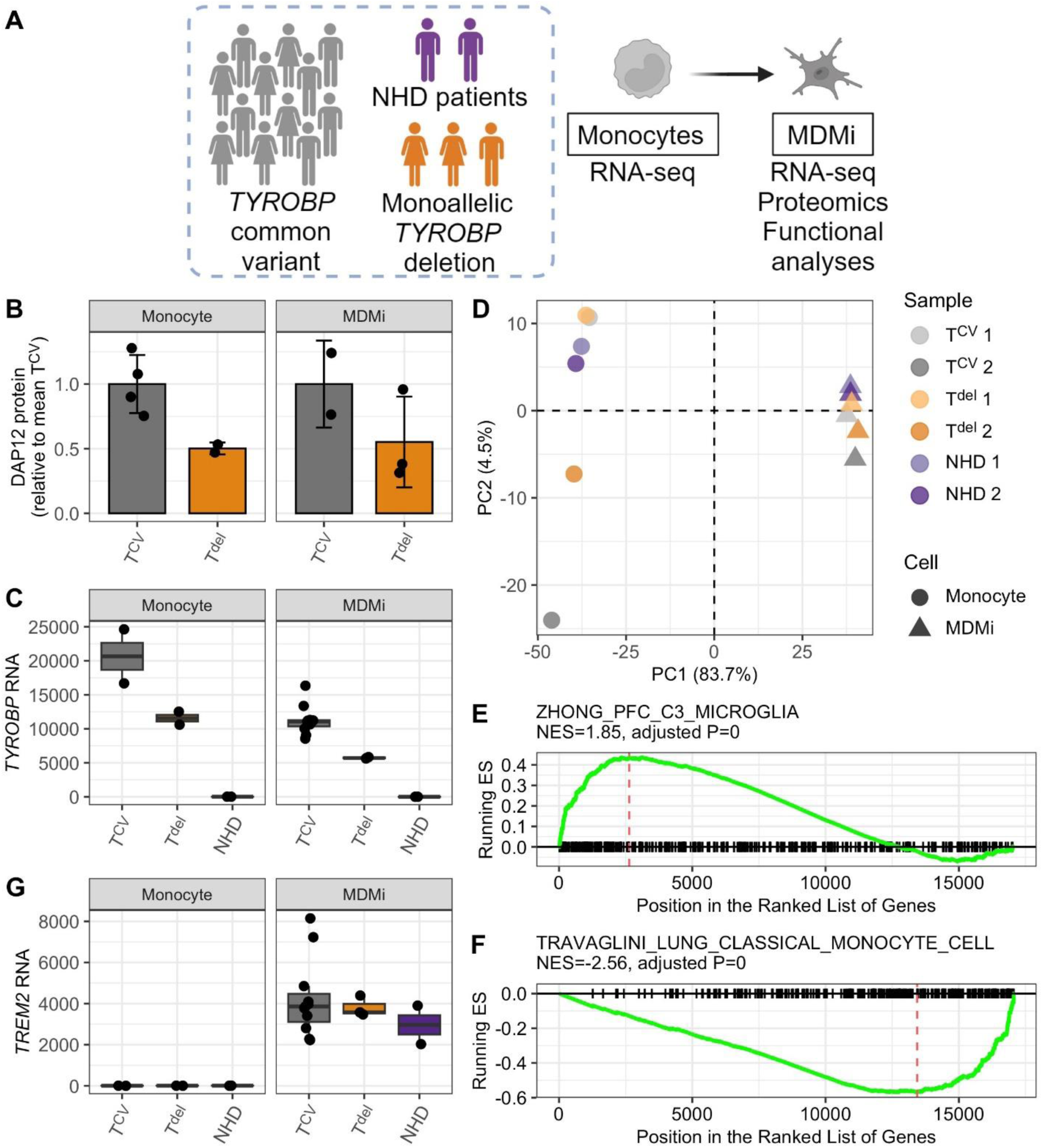
Characterization of the MDMi cell model. **(A)** Study design to assess functional effects of the *TYROBP* deletion in MDMi cells. **(B)** A non-significant but clear trend of decreased DAP12 protein levels in the monocytes and MDMi of monoallelic *TYROBP* deletion carriers compared to cells from individuals homozygous for the *TYROBP* common variant. DAP12 protein levels were normalized to GAPDH levels in the same lysate. Monocytes: T^CV^, n=4; T^del^, n=2; MDMi: T^CV^, n=2; T^del^, n=3. **(C)** *TYROBP* RNA levels are decreased in monocytes and MDMi of monoallelic *TYROBP* deletion carriers compared to cells from individuals homozygous for *TYROBP* common variant, while no *TYROBP* RNA is detected in NHD patients homozygous for the *TYROBP* deletion. Monocytes: T^CV^, n=2; T^del^, n=2; NHD, n=2; MDMi: T^CV^, n=8; T^del^, n=3; NHD, n=2. **(D)** Principal component analysis suggests successful differentiation from monocytes to MDMi in all genotype groups. **(E-F)** GSEA analysis shows that microglial genes are upregulated (E) while monocyte genes are downregulated (F) in the MDMi cells compared to monocytes. **(G)** *TREM2* is not expressed in the monocytes and is upregulated during MDMi differentiation in all genotype groups. Monocytes: T^CV^, n=2; T^del^, n=2; NHD, n=2; MDMi: T^CV^, n=8; T^del^, n=3; NHD, n=2. Data in B, C, and G are shown as mean ±SD. Each data point represents one individual.

To evaluate the success of microglia-like differentiation, global RNA-sequencing of monocytes and MDMi was carried out (Table S3). MDMi showed a distinct gene expression profile compared to monocytes as indicated by principal component analysis (PCA) which showed a robust clustering by the cell type regardless of the *TYROBP* genotype (Fig. 5D). Gene set enrichment analysis (GSEA) further confirmed upregulation of microglial genes (Fig. 5E) and downregulation of monocyte genes (Fig. 5F) in the MDMi. *TREM2*, the key microglial gene and receptor associated with DAP12, was expressed in the MDMi but not in monocytes and was not affected by the *TYROBP* genotype (Fig. 5G). These results indicate a robust differentiation of the monocytes into microglia-like MDMi cells independently of the *TYROBP* deletion genotype. Thus, MDMi are a valid model to study the effects of *TYROBP* deletion in microglia-like cells.

### *TYROBP* deletion induces changes in the inflammatory response, unfolded protein response, and cellular metabolic pathways in MDMi cells

To examine microglial effects of *TYROBP* deletion, we carried out differentially expressed gene (DEG) and protein (DEP) analyses in the MDMi cells derived from NHD patients, monoallelic *TYROBP* deletion carriers and individuals homozygous for *TYROBP* common variant. Myelin debris and LPS challenge for 24 h were used to mimic different CNS stress conditions and to activate the TREM2 and TLR4-mediated pathways, respectively. Monoallelic *TYROBP* deletion induced only two DEGs in untreated and myelin-treated conditions when compared to *TYROBP* common variant carriers (Fig. 6A-B, Tables S4-S5). In contrast, in the LPS-treated condition, several DEGs were observed (Fig.6C, Table S6). A similar trend was observed in the number of differentially expressed proteins (Fig. 6D-F, Tables S7-S9). Biallelic *TYROBP* deletion had a more robust effect on the gene and protein expression in all conditions when compared to individuals homozygous for the *TYROBP* common variant (Fig. S3A-E, Tables S10-S14) or monoallelic *TYROBP* deletion carriers (Fig. S4A-E, Tables S15-S19). As expected, *TYROBP* was not detected in the NHD MDMi at either the transcript or protein level, whereas a well-established NHD microglial marker CD163 was significantly upregulated at both transcript and protein levels in NHD MDMi when compared to individuals homozygous for *TYROBP* common variant in all comparisons, except in the DEG analysis of untreated cells (Fig. S3A-E).

**Figure 6.**
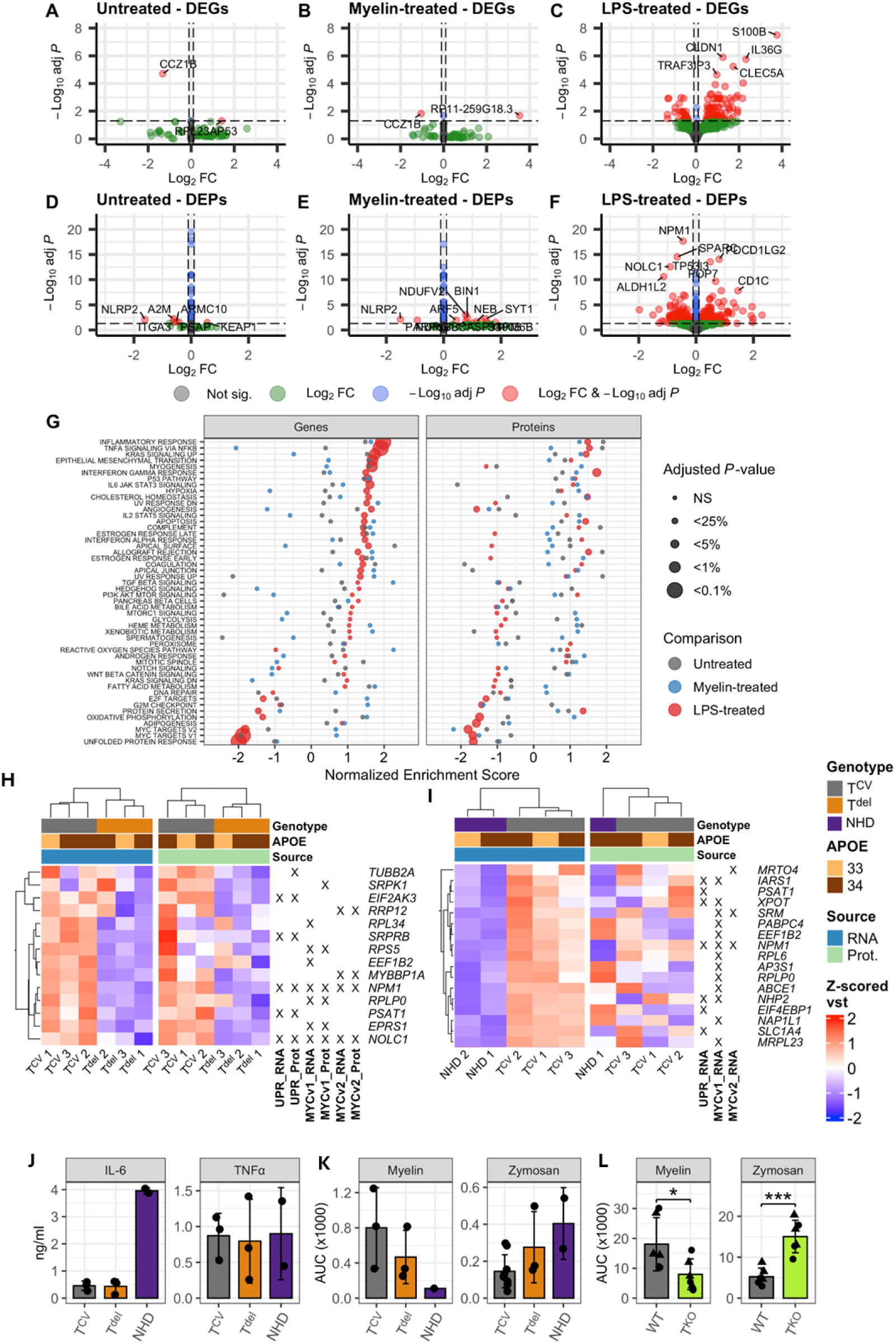
Increased inflammatory response and decreased unfolded protein response induced by monoallelic *TYROBP* deletion in MDMi cells upon LPS treatment. **(A-C)** Volcano plots of differentially expressed genes and **(D-F)** proteins in the monoallelic *TYROBP* deletion carrier MDMi cells compared to controls upon untreated (A, D), myelin-treated (B, E) and lipopolysaccharide (LPS)-treated (C, F) conditions., T^CV^, n=3-12; T^del^, n=3. **(G)** Pathway enrichment of genes (left panel) and proteins (right panel) differentially expressed in the monoallelic *TYROBP* deletion carrier MDMi cells compared to *TYROBP* common variant MDMi cells. **(H-I)** Heat maps showing the top up- and downregulated targets in the UPR and MYC pathways in the monoallelic (H) and biallelic (I) *TYROBP* deletion carriers compared to controls. **(J)** Inflammatory response was assessed by measuring interleukin-6 (IL-6), and tumor necrosis factor α (TNFα) levels in the conditioned media of T^CV^, T^del^, and NHD MDMi upon 24h LPS treatment. Each data point indicates one individual, an average of 1-3 replicate wells. T^CV^, n=3; T^del^, n=3; NHD, n=2. **(K)** Phagocytic activity of T^CV^, T^del^, and NHD MDMi was assessed by challenging the cells with pHrodo-labelled myelin or zymosan. Images were obtained every 15 min for a total of 3 hours. pHrodo positive area was quantified and normalized to cell count. Data are represented as an area under the curve (AUC) of 3-h phagocytic challenge. Each data point indicates one individual, an average of 2-3 technical replicates. T^CV^, n=3-6; T^del^, n=3; NHD, n=1-2. **(L)** Phagocytic activity in BV2 cells was assessed by quantifying the area of pHrodo-labelled myelin and zymosan signal normalized to confluency. Data are represented as AUC of 6-h phagocytic challenge. Each data point indicates an average of three technical replicates; triangles and circles represent two separate control and knockout (KO) lines, data pooled from three independent experiments. Independent samples T-test, *: *P*<0.05, ***: *P*<0.001. All data in J-L are shown as mean ± SD. T^CV^, *TYROBP* common variant; T^del^, monoallelic *TYROBP* deletion; T^KO^, *Tyrobp* knock out; NHD, Nasu-Hakola disease; WT, wild type.

Gene ontology enrichment was carried out to gain insight into the altered cellular processes induced by mono- or biallelic *TYROBP* deletion. Monoallelic *TYROBP* deletion induced significant upregulation of the inflammatory response, TNFα signaling via NFKB, KRAS signaling up, interferon gamma response, cholesterol homeostasis, apoptosis, and allograft rejection pathways both at the transcript and protein levels upon LPS treatment (Fig. 6G). Simultaneously, a significant downregulation of the unfolded protein response (UPR) and myc targets pathways was observed at both transcript and protein levels (Fig. 6G). Closer inspection of the downregulated pathways revealed, among others, reduced transcript and protein levels of *EIF2AK3*, which encodes PERK, one of the three sensors of UPR, suggesting that specifically the PERK arm of UPR might be downregulated in monoallelic *TYROBP* deletion carriers (Fig. 6H). No significant pathway enrichment was observed in untreated, or myelin-treated MDMi from monoallelic *TYROBP* deletion carriers when compared to individuals homozygous for *TYROBP* common variant.

Multiple inflammation related gene sets were upregulated in the NHD compared to *TYROBP* common variant or monoallelic *TYROBP* deletion MDMi, including interferon gamma response, TNFα signaling via NFKB, IL2-STAT5 signaling, and IL6-JAK-STAT3 signaling (Fig. S3F, Fig. S4F), which were also significantly upregulated in the NHD brain microglia according to a recent single nucleus RNA-sequencign (sn-RNAseq) analysis (*22*). Notably, these gene sets were upregulated in the NHD MDMi in all treatment conditions, suggesting a strong proinflammatory transcriptional profile in these cells even without an external stimulus. While the UPR was not among the top downregulated pathways in NHD MDMi when compared to controls, we explored it in more detail to assess potential similarities and differences in MDMi derived from monoallelic *TYROBP* deletion carriers and NHD patients in comparison to controls. Two common downregulated targets, PSAT1 and NPM1, were detected in this pathway (Fig. 6I).

To further validate the inflammatory phenotype induced by *TYROBP* deletion, secretion of IL-6, TNFα and IL-10 was measured in the conditioned medium collected from the MDMi cells after 24-h LPS stimulation. Approximately 9-fold increase in IL-6 secretion was detected in the NHD MDMi when compared to MDMi from monoallelic *TYROBP* deletion carriers or individuals homozygous for *TYROBP* common variant, while the TNFα levels did not differ between the genotypes (Fig. 6J). IL-10 was only detectable in the NHD samples, suggesting a marked increase in secretion of IL-10 in response to LPS, in line with the previous sn-RNAseq analysis in NHD brain microglia (*22*).

To further investigate the effects of *TYROBP* deletion on key microglia-related functions, pHrodo-labelled myelin and zymosan were applied to MDMi cells, and their phagocytic uptake was followed by live-cell imaging. A trend towards reduced myelin uptake was observed in mono- and biallelic *TYROBP* deletion carrying cells, as indicated by the smaller area of fluorescence signal in area under the curve (AUC) analysis (Fig. 6K, Fig. S5A). Opposed to that, the phagocytosis of zymosan appeared to be more active in mono- and biallelic *TYROBP* deletion carrying cells (Fig. 6K, Fig. S5B), however, the results were not statistically significant. To further validate these findings, *Tyrobp* knockout (KO) BV2 microglial cell lines were generated by lentiviral CRISPR-Cas9 editing (Fig. S6A-B), and the phagocytosis experiments were replicated in two KO lines and two control lines. The phagocytosis of myelin was significantly reduced in KO cells compared to control cells (*P*=0.0367, Independent samples T-test, Fig. 6L, Fig. S6C), whereas the uptake of zymosan was clearly higher in KO *vs*. control cells (*P*<0.001, Independent samples T-test, Fig. 6L, Fig. S6D). These findings indicate an expected loss in TREM2-mediated phagocytosis of myelin in the absence of DAP12, whereas the phagocytosis of zymosan is activated.

## DISCUSSION

In this study we show that monoallelic 5.2-kb *TYROBP* deletion is a novel risk factor for AD, leading to twofold increased risk of AD and dementia with an earlier onset age in the monoallelic carriers when compared to non-carriers. Furthermore, we observed that monoallelic *TYROBP* deletion induces cystic bone lesions typical to NHD caused by biallelic *TYROBP* or *TREM2* loss. The monoallelic *TYROBP* deletion leads to reduced levels of DAP12 protein (encoded by *TYROBP*) in myeloid cells, suggesting that reduction in DAP12 protein levels is the primary mechanism behind these effects, potentially leading to reduced signaling from DAP12-associated receptors. We used MDMi derived from peripheral blood monocytes from the carriers as a surrogate model for human CNS microglia, the only brain cell type that expresses *TYROBP*, to study the effects of *TYROBP* deletion. Our findings indicate that MDMi derived from monoallelic *TYROBP* deletion carriers display upregulated inflammatory response and downregulated unfolded protein response pathways upon LPS stimulation as compared to control MDMis.

To our knowledge, this is the first report to show genetic association at genome-wide significant level between the *TYROBP* locus and AD risk. *TYROBP* locus has not been pinpointed in any previous genome-wide association studies for AD, while targeted investigations have led to conflicting results. Enrichment of rare coding variants in *TYROBP* was detected in a cohort of early-onset AD patients (*12*), while no predicted pathogenic *TYROBP* variants were detected in a cohort of Turkish dementia patients (*10*). A study among 3,220 older Finns did not detect any association between monoallelic *TYROBP* deletion and cognitive impairment, and reported very low or absent Aβ pathology in the neocortex of two autopsied *TYROBP* deletion carriers (*11*). Due to the rarity of *TYROBP* deletion, only 11 monoallelic carriers were identified in this previous study, potentially limiting its power to detect associations. To overcome this limitation, we utilized FinnGen, the largest available Finnish cohort with genotype and health registry data on 520,210 individuals and detected 2,231 putative monoallelic *TYROBP* deletion carriers using a proxy marker. This approach allowed us to detect increased risk and earlier age at onset for AD and dementia among the monoallelic *TYROBP* deletion carriers compared to noncarriers.

While earlier genetic evidence has been inconclusive, network based analyses have identified *TYROBP* as a central hub in networks regulating AD pathology and microglial sensory functions (*1, 2*), highlighting the potential role of *TYROBP* in AD. Paradoxically, studies in mouse models have suggested that partial or complete loss of Tyrobp helps to normalize learning behavior deficits and electrophysiological properties associated to cerebral Aβ amyloidosis (*23, 24*) and tauopathy (*25*). Consequently, it has been postulated that reduction of DAP12 levels might present a therapeutic opportunity in AD. This starkly contrasts with our present findings in humans indicating that reduced DAP12 levels caused by monoallelic *TYROBP* loss significantly increase the risk and lower the age of onset in AD. These contradictory findings might arise from species-specific differences (*22*) or the fact that the AD mouse models cannot fully recapitulate the complexity of the human disease. Nevertheless, they emphasize the importance of human data and humanized models in the search for therapeutic targets in AD.

To begin to understand the functional effects of monoallelic *TYROBP* deletion in human microglial cells we used global transcriptomic and proteomic analyses in a monocyte-derived microglial cell model from the carriers. We found that 24-h LPS stimulation induces upregulation of inflammatory pathways and downregulation of MYC and UPR pathways in the monoallelic *TYROBP* deletion carrier MDMi as compared to MDMi derived from control individuals. Among the top downregulated targets, we highlight PERK (encoded by *EIF2AK3*). This is one of the three kinases that initiate the UPR signaling. Another identified target was PSAT1, a phosphoserine aminotransferase, whose levels were reduced at both transcript and protein levels in the MDMi from monoallelic *TYROBP* deletion carriers. Intriguingly, emerging results suggest that PERK and PSAT1 converge on metabolic reprogramming that is needed for promoting the immunosuppressive function in M2 macrophages (*26*) and differentiation of osteoclasts (*27*). Specifically, the studies suggest that PERK signaling mediates the upregulation of PSAT1 leading to increased production of α-ketoglutarate, which is an essential cofactor for JMJD3-mediated histone demethylation. This epigenetic mechanism controls genes related to immunosuppression in M2 macrophages and early stages of osteoclastogenesis. Accordingly, this notion unites the two seemingly unconnected pathological processes, bone abnormalities and increased microglial immune response, which we observed in the monoallelic *TYROBP* deletion carriers and the microglial cell model.

Cystic bone lesions accompanied by pain, swelling and pathological fractures in the ankles and wrists are the hallmark of the osseous stage of NHD caused by the loss of *TYROBP* or *TREM2* and precede the neurological symptoms. Remarkably, we show that similar cystic-like bone lesions can be detected in the monoallelic carriers of *TYROBP* deletion, corroborating an earlier observation in monoallelic siblings of an Austrian NHD patient carrying different *TYROBP* variants (*28*). Intriguingly, genetic silencing or pharmacological inhibition of PSAT1 in mouse osteoclasts leads to an impairment in osteoclast multinucleation (*27*), similar to that observed in monocyte-derived osteoclasts of NHD patients (*29*). Interestingly, some deleterious biallelic *TREM2* variants lead to frontotemporal dementia without bone involvement (*6, 7*). Together, this evidence suggests that DAP12 encoded by *TYROBP* may be more critical for the osteoclast function than TREM2.

One of the functions of TREM2-DAP12 signaling in microglia is to promote anti-inflammatory signaling (*30*). Thus, the upregulation of immune response pathways observed in monoallelic *TYROBP* deletion carrier and NHD MDMis might be due to the inability of these cells to promote anti-inflammatory signaling. Consequently, these cells may be unable to resolve the immune response once it is activated, similar to the inability of PERK or PSAT1-deficient macrophages to initiate immunosuppressive activity (*26*). A recent sn-RNA-seq analysis in *post-mortem* brain of NHD patients observed microglial signature related to wound healing driven by STAT3, RUNX1, and TGFβ (*22*). While we did not detect an exactly similar signature in our MDMi cell model, many of the enriched pathways related to immune response are the same in the NHD patient-derived MDMi and the microglia in *post-mortem* NHD brain. The differences may be due to that the *post-mortem* microglia represent the end stage of the disease, while the MDMi derived from patients in the early neurological stage and cultured *in vitro* may resemble the microglia in the earlier stages of the disease.

Our study has some limitations. We have used an imputed proxy marker to identify *TYROBP* deletion carriers in the FinnGen data since data on large structural variants is not available in FinnGen. It is possible that the use of a proxy marker leads to both inclusion of non-carriers (false positives) and exclusion of true deletion carriers (false negatives). However, we confirmed in a subcohort of 50 individuals from FinnGen by Sanger sequencing that the imputed proxy marker carriers are true 5.2-kb *TYROBP* deletion carriers, except for a rare case of very low imputation Info score, i.e. low quality of the imputed genotype, suggesting that our proxy marker reliably identifies the *TYROBP* deletion carriers. While loss of *TYROBP* expression is the most plausible consequence of the 5.2-kb deletion, we cannot exclude the possibility that the deletion has additional effects besides reduced DAP12 protein levels in myeloid lineage cells demonstrated in our study. While the deleted sequence does not contain any other coding regions besides the exons 1-4 of *TYROBP*, it is possible that the region contains regulatory elements, and their absence might affect the expression of other genes. Replication of our genetic association finding in other populations or additional cohorts is challenging since the 5.2 kb deletion of *TYROBP* is a Finnish population specific founder mutation, and our study was conducted using the largest available data release of the unique FinnGen study, combining genotype data and longitudinal health registry data of more than 500,000 individuals, covering approximately 10 % of the Finnish population. Other NHD causative *TYROBP* variants are known globally, especially in the Japanese population. However, they are extremely rare, and thus, finding enough monoallelic carriers in a well characterized cohort is unlikely. Nevertheless, our finding is logical due to the close functional connection between DAP12 (encoded by *TYROBP*) and TREM2, and the association of *TREM2* variants with AD risk. While our genetic association finding is robust, a limitation of the study is that we are only beginning to understand the functional effects of the monoallelic *TYROBP* deletion by using a monocyte-derived microglial cell model from a small number of donors. However, this is the first report exploring the functional effect of the monoallelic *TYROBP* deletion in microglial cells. Our key findings related to altered genes and pathways are supported by both transcriptomic and proteomic analysis.

In summary, this study identified the monoallelic 5.2-kb *TYROBP* deletion as a novel risk factor for AD and dementia, demonstrated for the first time NHD-like bone cysts in a monoallelic *TYROBP* deletion carrier, and provided insights into key biological pathways altered in microglial cells due to *TYROBP* deletion.

## MATERIALS AND METHODS

### Study design

The overall aim of our study was to assess the phenotypic and functional effects of a Finnish founder mutation, 5.2 kb *TYROBP* deletion, that is known to cause the early-onset neurodegenerative disease NHD in biallelic state. We hypothesized that the monoallelic *TYROBP* deletion may be a risk factor for neurodegeneration later in life and may induce functional changes in the microglial cells. In the first part of the study, we aimed to elucidate the clinical phenotype of the monoallelic *TYROBP* deletion in a large, well-characterized biobank cohort FinnGen. To achieve this aim, we first identified the founder haplotype and genetic proxy markers for the Finnish *TYROBP* deletion by utilizing WGS of three Finnish NHD patients.

Using the proxy markers, we conducted phenome-wide association study in FinnGen data freeze 12 (DF12) which contains genome and digital healthcare data on 520,210 Finnish individuals and 2489 clinical endpoints. To complement the picture of clinical phenotype induced by *TYROBP* deletion, we present case reports of two monoallelic *TYROBP* deletion carriers. The first case is a female in their third decade of life presenting classic NHD bone cysts in wrists and ankles. The second case is a female in their seventh decade of life diagnosed with idiopathic NPH (iNPH), with Aβ-positive frontal cortical brain biopsy and CSF biomarker profile indicative of AD-related brain pathology.

In the second part of the study, we aimed to experimentally evaluate the microglial effects of *TYROBP* deletion using *in vitro* models. Human monocytes for MDMi differentiation were extracted from peripheral venous blood samples obtained from three carriers of the Finnish *TYROBP* deletion (>60 years), 13 non-carrier age-matched controls, and two NHD patients (30-40 years). MDMi cultures in basal conditions or after treatment with myelin or LPS were used for omics and targeted functional analyses as detailed below. To validate the functional effects of *TYROBP* loss-of-function on microglial phagocytosis, we created *Tyrobp* KO mouse microglial BV2 cell lines. CRISPR-Cas9 genome editing was used to target mouse *Tyrobp*, and clonal selection was used to isolate monoclonal cell lines. Two *Tyrobp* KO lines and two control lines were used for subsequent experiments in basal conditions or after treatment with myelin or zymosan as detailed below.

All study protocols concerning human samples were approved by Medical Research Ethics Committee of Wellbeing Services County of North Savo (formerly Medical Research Ethics Committee of North Savo Hospital District). Written informed consent was obtained from all participants. The Ethics Committee of the Hospital District of Helsinki and Uusimaa (HUS) has coordinated the approval for FinnGen Study.

### Study subjects

NHD patients, monoallelic *TYROBP* deletion carriers, and unrelated controls were recruited from Neurology clinics at Kuopio University Hospital and Oulu University Hospital, or Kuopio University Hospital NPH registry (*19*), Finnish Geriatric Intervention Study to Prevent Cognitive Decline and Disability (FINGER) (*31*), and Auria biobank cohort during 2020-2023. Blood samples for monocyte and/or DNA isolation were collected following written informed consent from each participant.

The FinnGen Study (https://www.finngen.fi/en) is a large biobank-scale research project which combines genome data with digital healthcare data based on national health registers (*14*). FinnGen includes samples collected by the Finnish biobanks as well as legacy samples from previous research cohorts that have been transferred to the biobanks. FinnGen Study approved the use of the data in the present work.

The study subjects in FinnGen have provided informed consent for biobank research based on the Finnish Biobank Act. Alternatively, separate research cohorts that were collected prior to the Finnish Biobank Act coming into effect (September 2013) and start of FinnGen (August 2017), were collected based on study-specific consents and later transferred to the Finnish biobanks after approval by the Finnish Medicines Agency Fimea. Participant recruitment followed the biobank protocols approved by Fimea. The Coordinating Ethics Committee of the Hospital District of Helsinki and Uusimaa (HUS) statement number for the FinnGen study is HUS/990/2017. The complete list of ethics committee approval numbers, study permits, and biobank sample and data accession numbers are included in the Supplementary material.

### Genotyping

For non-FinnGen study participants, genomic DNA was extracted from peripheral whole blood using QIAamp DNA Blood Mini Kit (Qiagen, Hilden, Germany). DNA for a subset of 50 imputed *TYROBP* deletion proxy marker carriers from the FinnGen cohort was obtained through the Biobank of Eastern Finland.

Library preparation and WGS on Illumina NovaSeq sequencing platform was carried out at Novogene (Novogene (UK) Company Limited, Cambridge, UK). WGS data were initially processed using the nf-core sarek pipeline (release 3.1) with default settings (*32*), and the GATK GRCh38 as the reference genome. After identifying potential problems in the initial read alignment at and near the deletion breakpoints, WGS data were aligned again to the human reference genome (GRCh38) using a splice-aware aligner STAR (v2.7.9a) (*33*) with essential non-default settings: --outFilterMultimapNmax 1, --outFilterMismatchNmax 3, --alignIntronMax 10000 and --alignMatesGapMax 10000.

*TYROBP* deletion specific PCR was carried out as described previously (*11*). Genotyping of SNV 19:35901079-T-G was carried out by Sanger sequencing. In brief, the target region was amplified by PCR using primers 5’-GCGAACGCAGTCCCTGAATGG-3’ (forward) and 5’-CCTCCCTCTGGACCCAGTAA-3’ (reverse) and the PCR product was cleaned using NucleoSpin Gel and PCR Clean-up mini kit (Macherey-Nagel, Düren, Germany). The purified PCR product was combined with the reverse primer and sent to Macrogen Europe (Amsterdam, the Netherlands) for Sanger sequencing. Sanger sequencing was reliable only from reverse direction due to the presence of several poly-T repeats between the forward primer and the variant. *APOE* genotyping was carried out with pre-designed TaqMan SNP genotyping assays for rs429358 and rs7412 (both from ThermoFisher Scientific). TaqMan SNP genotyping assays were performed according to manufacturer’s instructions, and all samples were assayed in duplicate.

The whole FinnGen cohort has been genotyped with multiple Illumina (Illumina Inc., San Diego, USA) and Affymetrix (Thermo Fisher Scientific, Santa Clara, CA, USA) chip arrays as part of the FinnGen Study. Chip genotype data were imputed using the Finnish population-specific imputation reference panel Sequencing Initiative Suomi project (SISu v4.2), Institute for Molecular Medicine Finland (FIMM), University of Helsinki, Finland (http://sisuproject.fi).

### FinnGen analyses

Phenotype information and clinical endpoints in FinnGen are based on different national health registries, including hospital discharge registers, prescription medication purchase registers, and cancer registers. A complete list of FinnGen endpoints and their respective controls are available at https://www.finngen.fi/en/researchers/clinical-endpoints and can be explored at https://risteys.finngen.fi/.

In this study we used summary statistics and data from the FinnGen data release R12. GWAS studies for FinnGen core endpoints were performed with REGENIE 2.2.4. A detailed description of the analytical methods is available at https://github.com/FINNGEN/regenie-pipelines. Phenotype associations for variant rs1244787406-G across 2489 FinnGen core analysis endpoints were visualized using LAVAA (*34*). Regional association plots for the endpoint ‘dementia including primary care registry’ were generated with topr 2.0.0 package in R 4.3.2 (*35, 36*). Linkage disequilibrium in-sample dosage in the *TYROBP*-locus (3 Mb window around the lead variant) in FinnGen was computed using LDstore2 (*37*) and fine-mapping was carried out using SuSiE (*38*) with the maximum number of causal variants in a locus L = 10.

The map visualizing the regional allele frequency of rs1244787406-G was created in R based on region of birth for minor and major allele carriers. Kaplan-Meier curves were drawn in R using package survminer v0.4.9 (*39*).

### Radiological imaging

The monoallelic *TYROBP* deletion carrier and the NHD patient were imaged as part of diagnostic procedure with conventional x-rays (Siemens Ysio Max, Erlangen, Germany) for skeletal features of the hands and feet, and at 3,0 Tesla MRI (Philips Achieva, Best, NL) and 1,5 Tesla MRI (GE Signa Artist, Milwaukee, USA) with standard clinical sequences, including T1, T2, FLAIR, DWI, and susceptibility weighted imaging, for potential brain pathology. The 3D T1 MRI data were also analyzed by brain volumetry software, cNeuro (Combinostics Ltd, Tampere, Finland).

### Immunohistochemistry and CSF biomarkers

Diagnostic brain biopsy specimen collected during NPH shunt surgery were used for immunohistochemical analysis. Immunostaining for Iba-1 and Aβ was performed as described previously (*40*). Full section brightfield images were obtained with Hamamatsu NanoZoomer-XR Digital slide scanner with 20x (NA 0.75) objective (Hamamatsu Photonics K.K., Shizuoka, Japan) and analyzed as described previously (*40*). In short, Aβ plaques were manually outlined in NDP.view2 software (Hamamatsu Photonics K.K.) to obtain the plaque size. The plaque-associated microglia with clearly visible soma were manually counted by an investigator blinded to sample identity.

CSF samples from the same NPH patients were obtained by lumbar puncture. Levels of AD-related biomarkers Aβ42, T-Tau, and P-Tau181 were analyzed using a commercial enzyme-linked immunosorbent assay (Innotest, Fujirebio, Ghent, Belgium).

### MDMi differentiation, culture, and treatments

To extract peripheral blood mononuclear cells (PBMCs), 60-100 ml peripheral venous blood was collected from participants and processed within 24 h of sample collection. PBMCs were extracted by density gradient centrifugation over Ficoll-Paque PLUS (#17-1440-02, Cytiva, Marlborough, MA, USA) in SepMate-50 tubes (#85450 Stemcell Technologies, Vancouver, Canada). CD14-positive monocytes were selected from PBMCs using human CD14 MicroBeads (Miltenyi Biotec, Bergisch Gladbach, Germany) and magnetic-activated cell sorting.

Monocytes were plated at 0.5 x 10^6^ cells per well in 12-well plates (for RNA-sequencing, LC-MS/MS, alphaLISA and conditioned medium) or at 1 x 10^5^ cells per well in 96-well plates (for phagocytosis). Monocyte differentiation into MDMis was done according to a previously published protocol (*20*), by culturing the monocytes under standard humidified environment (+37°C, 5% CO2) in MDMi culture medium consisting of RPMI-1640 Glutamax (#61870036, Gibco, Billings, MT, USA) supplemented with 1 % penicillin/streptomycin and a mixture of the following human recombinant cytokines: M-CSF (10 ng/ml, #574804, Biolegend, San Diego, CA, USA), GM-CSF (10 ng/ml, #215-GM-010/CF, R&D Systems, Minneapolis, MN, USA), NGF-β (10 ng/ml, #256-GF-100, R&D Systems), CCL2 (100 ng/ml, #571404, Biolegend), and IL-34 (100 ng/ml, #5265-IL-010/CF, R&D Systems).

After MDMi differentiation for 10-12 days *in vitro* (DIV), conditioned medium was replaced with fresh MDMi culture medium or with culture medium containing myelin (25 µg/ml) or LPS (200 ng/ml, O26:B6, L5543, Sigma Aldrich), and the cells were cultured for 24 h prior to sample collection. Conditioned medium was collected, centrifuged at 5 000 x *g* at +4°C for 10 min and stored at -80°C until used for further analyses. Cells were washed thrice with ice-cold PBS and subjected to RNA or protein extraction. For total RNA extraction, cells from 2-3 replicate wells/donor were collected into ice-cold PBS and RNA was extracted immediately. For protein extraction for LC-MS and alphaLISA experiments, cells were lysed in RIPA supplemented with protease and phosphatase inhibitors (Halt™ 78420 and 87786, Thermo Fisher) or 1x alphaLISA lysis buffer (PerkinElmer), respectively, and stored at -80°C until used for further analyses.

### Creation and validation of *Tyrobp* KO BV2 cell lines

Immortalized mouse microglial BV2 cells were cultured in RPMI-1640 medium (R0883-500ML, Sigma-Aldrich) supplemented with 10 % fetal bovine serum (10500064, Thermo Fisher), 1 % penicillin-streptomycin (DE17-602E, Lonza™), and 2 mM L-glutamine (BE17-605E, Thermo Fisher) in humidified atmosphere in 37°C with 5 % CO2. For the experiments, the concentration of FBS was lowered to 5%.

To generate *Tyrobp* KO cell lines, BV2 cells were transduced with LV01 all-in-one CRISPR-Cas9 lentiviral vector targeting exon three of *Tyrobp* (MMPD0000041569, Sigma-Aldrich). To generate control lines, LV01 non-targeting NegativeControl1 vector was used.

A total of 600,000 cells were seeded on 6-well plates and let to adhere for 2 h before adding the viral vector at multiplicity of infection (MOI) 10 together with 8 µg/ml of polybrene (TR-1003-G, Sigma-Aldrich). After 21 hours, the virus containing medium was replaced with fresh medium, and the puromycin selection (3 µg/ml, P8833-10MG, Sigma-Aldrich) was started four hours later. After four days of selection, monoclonal lines were established by sorting single GFP-positive cells on 96-wells by fluorescence-activated cell sorting. Monoclonal cell populations were expanded, and DNA and protein samples were collected for determining the knock-out lines. Sanger sequencing for *Tyrobp* exon three was performed to identify clones with successful gene editing.

The region was amplified by PCR using primers 5’-TTCTCCTTAGGATTAAGTCCCGT-3’ (forward) and 5’-TGTGACCTTGACGCTTCCAC-3’ (reverse), and the product was purified with ExoSAP-IT™ PCR Product Cleanup Reagent (78201.1.ML, Thermo Fisher Scientific). The sequencing reaction was performed using BigDye™ Terminator v3.1 kit (4337455, Thermo Fisher Scientific), and after ethanol/EDTA precipitation, the pure product was resuspended in Hi-Di™ Formamide (4311320, Thermo Fisher Scientific). The sequencing was performed at Genome Center of Eastern Finland. TIDE online tool (http://shinyapps.datacurators.nl/tide/) was used to investigate the generated indels in the lines.

DAP12 protein levels were investigated by Western Blotting from RIPA-extracted (89901, Thermo Fisher) protein samples as previously described (*40*) using the following primary and HRP-conjugated secondary antibodies: Recombinant rabbit anti-mouse DAP12 [EPR24244-119] (1:1000, ab280568, Abcam), mouse anti-β-actin (1:1000, ab8226, Abcam), secondary anti-rabbit-HRP (1:5000 NA934 Cytiva), and sheep anti-mouse-HRP (1:5000, NA931V, GE Healthcare, Chicago, IL, USA).

In total, seven clones had frameshift mutations in both copies of *Tyrobp* and no detectable DAP12 protein expression. Two knock-out clones (KO1 and KO2) were selected for the experiments, and two monoclonal lines transduced with the non-targeting vector were selected as control lines (CTRL1 and CTRL2). The proliferation of the selected lines was followed by IncuCyte S3 live cell imaging system, and no differences were detected in the growth rate of the lines in the time window of the performed experiments (Fig. S6B).

### RNA extraction and RNA-sequencing

Acutely isolated monocytes were placed in Macherey-Nagel™ NucleoProtect RNA reagent (ThermoFisher Scientific) and stored at +4°C until RNA extraction. MDMi collected in PBS as described above were subjected directly to RNA extraction. RNA was isolated using High Pure RNA Isolation Kit (11828665001, Roche), and the RNA extracts were stored at -80°C until further use.

Library preparation and RNA sequencing was conducted by Novogene (UK) Company Limited. In brief, mRNA enrichment was performed with oligo(dT) bead pulldown, from where the pulldown material was subjected to fragmentation, followed by reverse transcription, second strand synthesis, A-tailing, and sequencing adaptor ligation. The final amplified and size selected library comprised 250-300-bp insert cDNA and paired-end 150 bp sequencing was executed with an Illumina high-throughput sequencing platform. Sequencing yielded 4.4–26.9 million sequenced fragments per sample.

The 150 nucleotide pair-end RNA-seq reads were quality-controlled using FastQC (version 0.11.7) (https://www.bioinformatics.babraham.ac.uk/projects/fastqc/). Reads were then trimmed with Trimmomatic (version 0.39) (*41*) to remove Illumina sequencing adapters and poor quality read ends, using as essential settings: ILLUMINACLIP:2:30:10:2:true, SLIDINGWINDOW:4:10, LEADING:3, TRAILING:3, MINLEN:50. Trimmed reads were aligned to the Gencode human transcriptome version 38 (for genome version hg38) using STAR (version 2.7.9a) (*33*) with essential non-default settings: --seedSearchStartLmax 12, -- alignSJoverhangMin 15, --outFilterMultimapNmax 100, --outFilterMismatchNmax 33, -- outFilterMatchNminOverLread 0, --outFilterScoreMinOverLread 0.3, and --outFilterType BySJout. The unstranded, uniquely mapping, gene-wise counts for primary alignments produced by STAR were collected in R (version 4.2.2) using Rsubread::featureCounts (version 2.12.3) (*42*), ranging from 3.3 to 22.1 million per sample. Differentially expressed genes (DEGs) between experimental groups were identified in R (version 4.2.0) using DESeq2 (version 1.36.0) (*43*) by employing Wald statistic and lfcShrink for FC shrinkage (type=“apeglm”) (*44*).

Comparisons between TYROBP genotype groups were performed adjusting for the *APOE* ε genotype, and between monocytes and MDMi cells by adjusting for donor (equivalent of paired test). Pathway enrichment analysis was performed on the gene lists ranked by the pairwise DEG test log2FCs in R using clusterProfiler::GSEA (version 4.4.4) (*45*) with Molecular Signatures Database gene sets (MSigDB, version 2022.1) (*46*).

### Proteomics analysis

Cultured MDMi were lysed in RIPA lysis buffer (50 mM Tris-Cl, 150 mM NaCl, 1 % NP-40, 0.05 % sodium deoxycholate, 0.01 % SDS, pH 7.5) by incubation on ice for 20 min with intermediate vortexing. Cell debris and undissolved material were removed by centrifugation at 16.000 × g for 10 min at 4 °C, and protein concentration was determined using a BCA Assay (Thermo Fisher Scientific). 10 µg of each sample were diluted 1:2 in water and benzonase (Sigma-Aldrich) digestion was performed with 10 units for 30 min at 37 °C to remove remaining DNA/RNA. Protein digestion was performed with 125 ng LysC and 125 µg trypsin (Promega) using the single-pot solid-phase enhanced sample preparation (SP3) (*47*). The peptide solution was filtered through 0.22 µm Costar SPIN-X columns (Corning) and dried by vacuum centrifugation. Samples were dissolved in 20 µL 0.1% formic acid (FA) using a sonication batch (Hielscher) and the peptide concentration was measured using the Qubit protein assay (Thermo Fisher Scientific).

An amount of 350 ng of peptides were separated on a on an in-house packed C18 analytical column (15 cm × 75 µm ID, ReproSil-Pur 120 C18-AQ, 1.9 µm, Dr. Maisch GmbH) using a binary gradient of water and acetonitrile (B) containing 0.1% formic acid at flow rate of 300 nL/min (0 min, 2% B; 2 min, 5% B; 70 min, 24% B; 85 min, 35% B; 90 min, 60% B) and a column temperature of 50°C. A Data Independent Acquisition Parallel Accumulation–Serial Fragmentation (DIA-PASEF) method with a cycle time of 1.4 s was used for spectrum acquisition. Briefly, ion accumulation and separation using Trapped Ion Mobility Spectrometry (TIMS) was set to a ramp time of 100 ms. One scan cycle included one TIMS full MS scan and The DIA PASEF windows covered the m/z range from 350-1000 m/z with 26 windows of 27 m/z with an overlap of 1 m/z. The raw data was analyzed using the software DIA-NN version 1.8 (*48*) for protein label-free quantification (LFQ). A one protein per gene canonical fasta database of Homo Sapiens (download date March 1st 2023, 20603 entries) from UniProt and a fasta database with 246 common potential contaminations from Maxquant (*49*)were used to generate a spectral library in DIA-NN with a library free search which included 10615 proteins. Trypsin was defined as protease. Two missed cleavages were allowed, and peptide charge states were set to 2-4. Carbamidomethylation of cysteine was defined as static modification. Acetylation of the protein N-term as well as oxidation of methionine were set as variable modifications. The false discovery rate for both peptides and proteins was adjusted to less than 1 %. Data normalization was disabled.

Identification of differentially expressed, *APOE* ε genotype-corrected proteins between experimental groups, non-normalized LFQ intensities as the starting point, and the subsequent pathway enrichment were done as for the RNA-seq data, except using version 4.2.3 for R and version 1.38.3 for DESeq2 (*43*).

### AlphaLISA

PBMCs were extracted via density gradient centrifugation using Ficoll-Paque PLUS and stored in CryoStor® CS10 (07930, Stemcell Technologies) in liquid nitrogen. On the day of the monocyte isolation, one vial of PBMCs (approximately 5 million cells) from each donor was thawed and warmed up to 37°C before diluting the suspension with RPMI medium containing 20 % FBS. After centrifugation, the cells were resuspended in RPMI-FBS and incubated for 30 min at 37°C to allow the cells to recover from the thawing. CD14+ monocytes were isolated by magnetic-activated cell sorting using CD14 MicroBeads as described above, and the resulting monocyte fraction was lysed in AlphaLISA lysis buffer. MDMi were prepared from freshly isolated monocytes as described earlier and lysed in AlphaLISA lysis buffer. The lysates were stored at -20°C until measured.

Total DAP12 levels were measured from the cell lysates using AlphaLISA Surefire Ultra Total DAP12 Detection Kit (ALSU-TDAP12-A-HV, Revvity), and the results were normalized to GAPDH levels measured from the same lysate using AlphaLISA Surefire Ultra Human and Mouse Total GAPDH Detection Kit (ALSU-TGAPD-B-HV, Revvity).

### Cytokine ELISAs in conditioned medium

To study the inflammatory response in MDMi cells, IL-6, IL-10 and TNFα were measured from the conditioned medium using commercial ELISA kits (IL-6: 88-7066-22, IL-10: 88-7106-22, TNFα: 88-7346-22, Thermo Fisher) according to manufacturers instructions.

### Myelin isolation and labeling

Myelin was isolated from adult male C57BL/6J mice using the sucrose gradient protocol described elsewhere (*50, 51*). 0.85 M and 0.32 M sucrose solutions were used for the gradient and Beckman Ultracentrifuge with Ti 50.2 rotor for the centrifugation. The concentration of the myelin preparation was determined using Pierce BCA Protein Assay kit (23225, Thermo Scientific) and stored at -80°C until used. Myelin was labelled with pHrodo™ iFL Red Microscale Protein Labeling Kit (P36014, Thermo-Fisher Scientific). For labelling, 1 µl of dye was added per 40 µg of myelin and the mixture was incubated for one hour at room temperature. The excess dye was removed by washing twice with PBS, and after the final centrifugation (15,000 x *g*, 10 min), the myelin pellet was resuspended in PBS at 1 mg/ml and stored at -80°C until used.

### Phagocytosis assays

For phagocytosis analysis, MDMi and BV2 cell lines were challenged with myelin debris or zymosan bioparticles. Monocytes were plated at 50,000 cells per well in 96-well plates and differentiated into MDMi for 10-12 DIV. BV2 cells were plated at 6,000 cells per well in 96-well plates and let to adhere for 2 hours. Labelled myelin was applied to the cells at 25 µg/ml final concentration. Alternatively, pHrodo-zymosan bioparticles (Essen Bioscience) were applied at 50 µg/ml final concentration. Immediately following addition of the phagocytic targets, the culture plates were placed in IncuCyte S3 and imaged every 15 minutes for 3 h (MDMi) or 30 minutes for 6 hours (BV2) with red fluorescent and phase contrast channels using 10x and 20x objectives for MDMi and BV2 cells, respectively. Two to three replicate wells were used per sample donor or per BV2 cell line, and four fields of view were imaged per well. To allow normalization of the phagocytosis data by cell count, MDMi cell nuclei were stained with 1 µM Vybrant™ DyeCycle™ Green Stain (ThermoFisher Scientific) at the end of the 3 h assay, and imaged with the same image settings with the addition of green fluorescent channel. For BV2 cell normalization, a phase contrast image was captured before the addition of phagocytic targets, and the data were normalized to cell confluency. To validate the specificity of the phagocytosis signal, replicate wells without phagocytic targets, or with the actin polymerization inhibitor Cytochalasin D (10 µM; C8273-1MG, Sigma-Aldrich) which inhibits phagocytic uptake, were included in each phagocytosis experiment.

Segmentation of the red fluorescence signal was done using the IncuCyte Basic Analyzer and measured as area (µm^2^) per image at each time point. For MDMi cells, green fluorescent signal was segmented and object count per image was performed. Area under the curve of red fluorescent signal indicating phagocytic uptake was calculated from the normalized data in GraphPad Prism v10.

### Statistical analysis

Statistical analyses and data visualization were performed in R 4.3.2 (*36*) and GraphPad Prism v10. Chi-squared test was used to analyze regional allele frequency and *APOE* carrier frequency. Survival analysis comparing disease-free survival of rs1244787406-G carriers and noncarriers was performed in R where Kaplan-Meier curves were drawn using package survminer v0.4.9 (*39*) and log-rank test was performed with package survival v3.2-7 (*52, 53*). Independent samples T-test was used to analyze normally distributed immunohistochemistry and CSF biomarker data between the *APOE* ε3ε3 and ε3ε4 groups; cytokine and phagocytosis data between monoallelic *TYROBP* deletion carriers and noncarriers; and phagocytosis data between *Tyrobp* KO and control lines. Additional information on the statistical test, sample size, and technical replicates is given in the figure legends.

## Data Availability

All data associated with this study are present in the paper or the Supplementary Materials. The study participant consent does not allow opening the sequencing (WGS, RNA-seq) or proteomic data generated and analyzed during the current study, but they are available from the corresponding authors (H.M. or M.H.) on reasonable request. Summary statistics from each FinnGen data release will be made publicly available after a one-year embargo period and can be accessed freely at www.finngen.fi/en/access_results. For individual level data, the Finnish biobank data can be accessed through the Fingenious services (https://site.fingenious.fi/en/) managed by FINBB. Access to Finnish Health register data can be applied from Findata (https://findata.fi/en/data/).

## List of Supplementary Materials

Materials and methods Fig. S1 to S6

Tables S1 to S19

## Acknowledgments

We are thankful for the NHD patients and their families, and other sample donors for their participation in the study. We acknowledge Research Nurse Tanja Kumpulainen for her valuable work. The computational analyzes for WGS, RNA-sequencing, and proteome data were performed on servers provided by UEF Bioinformatics Center, University of Eastern Finland, Finland. Immunohistochemistry and live cell imaging with IncuCyte were carried out using infrastructure of UEF Cell and Tissue Imaging Unit, University of Eastern Finland, Biocenter Kuopio and Biocenter Finland. We acknowledge the participants and investigators of FinnGen study. Following biobanks are acknowledged for delivering biobank samples to FinnGen: Auria Biobank (www.auria.fi/biopankki), THL Biobank (www.thl.fi/biobank), Helsinki Biobank (www.helsinginbiopankki.fi), Biobank Borealis of Northern Finland (https://www.ppshp.fi/Tutkimus-ja-opetus/Biopankki/Pages/Biobank-Borealis-briefly-in-English.aspx), Finnish Clinical Biobank Tampere (www.tays.fi/en-US/Research_and_development/Finnish_Clinical_Biobank_Tampere), Biobank of Eastern Finland (www.ita-suomenbiopankki.fi/en), Central Finland Biobank (www.ksshp.fi/fi-FI/Potilaalle/Biopankki), Finnish Red Cross Blood Service Biobank (www.veripalvelu.fi/verenluovutus/biopankkitoiminta), Terveystalo Biobank (www.terveystalo.com/fi/Yritystietoa/Terveystalo-Biopankki/Biopankki/) and Arctic Biobank (https://www.oulu.fi/en/university/faculties-and-units/faculty-medicine/northern-finland-birth-cohorts-and-arctic-biobank). All Finnish Biobanks are members of BBMRI.fi infrastructure (www.bbmri.fi). Finnish Biobank Cooperative -FINBB (https://finbb.fi/) is the coordinator of BBMRI-ERIC operations in Finland.

## Funding

Research Council of Finland, grant 355604 (HM), grant 338182 (MH), grant 330178 (MT), grant 339767 (VL).

Faculty of Health Sciences, University of Eastern Finland (HM, MT).

Sigrid Jusélius Foundation (MH, VL, ES, TN).

The Strategic Neuroscience Funding of the University of Eastern Finland (MH, AH, VL, ES). Alzheimer’s Association ADSF-24-1284326-C (MH).

The State Research Funding KUH-VTR (VL, ES). Doctoral Programme in Molecular Medicine (RMW, HJ).

Deutsche Forschungsgemeinschaft (DFG, German Research Foundation) under Germany’s Excellence Strategy within the framework of the Munich Cluster for Systems Neurology, EXC 2145 SyNergy – ID 390857198 (CH and SFL) and a Koselleck Project HA1737/16-1 (CH).

JPco-fuND 2019 Personalised Medicine for Neurodegenerative Diseases; PMG-AD (CH, SFL, JCL, and MH; local funding agency grant numbers 01ED2002B and 334802).

The FinnGen project is funded by two grants from Business Finland (HUS 4685/31/2016 and UH 4386/31/2016) and the following industry partners: AbbVie Inc., AstraZeneca UK Ltd, Biogen MA Inc., Bristol Myers Squibb (and Celgene Corporation & Celgene International II Sàrl), Genentech Inc., Merck Sharp & Dohme LCC, Pfizer Inc., GlaxoSmithKline Intellectual Property Development Ltd., Sanofi US Services Inc., Maze Therapeutics Inc., Janssen Biotech Inc, Novartis Pharma AG, and Boehringer Ingelheim International GmbH.

## Author contributions

Conceptualization: HM, MH Methodology: HM, RMW, MT, PIM

Formal analysis: HM, RMW, SH, SAM, TK

Investigation: HM, RMW, SAM, RS, MT, PM, VP, AGR, PH, SPJ, HJ, IK, KS, MN, SVH, MIK, JM, JH, TR

Resources: JM, TN, JL, CB, JCL, CH, JR, JH, TR, JK, HS, AH, SFL, VL, ES, MH

Writing – original draft: HM

Writing – review & editing: HM, RMW, SH, SAM, MT, SPJ, IK, MN, SVH, MIK, TN, CB, JH, JK, AH, ES, MH

Visualization: HM, SH, RMW Supervision: HM, MH

Project administration: HM

Funding acquisition: HM, RMW, MT, HJ, TN, JCL, CH, AH, SFL, VL, ES, MH

## Competing interests

CH collaborates with Denali Therapeutics and is a member of the advisory boards of AviadoBio and Cure Ventures. The other authors declare that they have no competing interests.

## Data and materials availability

All data associated with this study are present in the paper or the Supplementary Materials. The study participant consent does not allow opening the sequencing (WGS, RNA-seq) or proteomic data generated and analyzed during the current study, but they are available from the corresponding authors (H.M. or M.H.) on reasonable request. Summary statistics from each FinnGen data release will be made publicly available after a one-year embargo period and can be accessed freely at www.finngen.fi/en/access_results. For individual level data, the Finnish biobank data can be accessed through the Fingenious® services (https://site.fingenious.fi/en/) managed by FINBB. Access to Finnish Health register data can be applied from Findata (https://findata.fi/en/data/).

## Consortia

### FinnGen

Aarno Palotie, Mark Daly, Bridget Riley-Gills, Howard Jacob, Coralie Viollet, Slavé Petrovski, Chia-Yen Chen, Sally John, George Okafo, Robert Plenge, Joseph Maranville, Mark McCarthy, Rion Pendergrass, Jonathan Davitte, Kirsi Auro, Simonne Longerich, Anders Mälarstig, Anna Vlahiotis, Katherine Klinger, Clement Chatelain, Matthias Gossel, Karol Estrada, Robert Graham, Dawn Waterworth, Chris ÓDonnell, Nicole Renaud, Tomi P. Mäkelä, Jaakko Kaprio, Minna Ruddock, Petri Virolainen, Antti Hakanen, Terhi Kilpi, Markus Perola, Jukka Partanen, Taneli Raivio, Jani Tikkanen, Raisa Serpi, Kati Kristiansson, Veli-Matti Kosma, Jari Laukkanen, Marco Hautalahti, Outi Tuovila, Jeffrey Waring, Bridget Riley-Gillis, Fedik Rahimov, Ioanna Tachmazidou, Zhihao Ding, Marc Jung, Hanati Tuoken, Shameek Biswas, Neha Raghavan, Adriana Huertas-Vazquez, Jae-Hoon Sul, Xinli Hu, Åsa Hedman, Máen Obeidat, Jonathan Chung, Jonas Zierer, Mari Niemi, Samuli Ripatti, Johanna Schleutker, Mikko Arvas, Olli Carpén, Reetta Hinttala, Johannes Kettunen, Arto Mannermaa, Katriina Aalto-Setälä, Mika Kähönen, Johanna Mäkelä, Reetta Kälviäinen, Valtteri Julkunen, Hilkka Soininen, Anne Remes, Mikko Hiltunen, Jukka Peltola, Minna Raivio, Pentti Tienari, Juha Rinne, Roosa Kallionpää, Juulia Partanen, Adam Ziemann, Nizar Smaoui, Anne Lehtonen, Susan Eaton, Heiko Runz, Sanni Lahdenperä, Natalie Bowers, Edmond Teng, Fanli Xu, Laura Addis, John Eicher, Qingqin S Li, Karen He, Ekaterina Khramtsova, Martti Färkkilä, Jukka Koskela, Sampsa Pikkarainen, Airi Jussila, Katri Kaukinen, Timo Blomster, Mikko Kiviniemi, Markku Voutilainen, Tim Lu, Linda McCarthy, Amy Hart, Meijian Guan, Jason Miller, Kirsi Kalpala, Melissa Miller, Kari Eklund, Antti Palomäki, Pia Isomäki, Laura Pirilä, Oili Kaipiainen-Seppänen, Johanna Huhtakangas, Nina Mars, Apinya Lertratanakul, Marla Hochfeld, Jorge Esparza Gordillo, Fabiana Farias, Nan Bing, Tarja Laitinen, Margit Pelkonen, Paula Kauppi, Hannu Kankaanranta, Terttu Harju, Riitta Lahesmaa, Hubert Chen, Joanna Betts, Rajashree Mishra, Majd Mouded, Debby Ngo, Teemu Niiranen, Felix Vaura, Veikko Salomaa, Kaj Metsärinne, Jenni Aittokallio, Jussi Hernesniemi, Daniel Gordin, Juha Sinisalo, Marja-Riitta Taskinen, Tiinamaija Tuomi, Timo Hiltunen, Amanda Elliott, Mary Pat Reeve, Sanni Ruotsalainen, Dirk Paul, Audrey Chu, Dermot Reilly, Mike Mendelson, Jaakko Parkkinen, Tuomo Meretoja, Heikki Joensuu, Johanna Mattson, Eveliina Salminen, Annika Auranen, Peeter Karihtala, Päivi Auvinen, Klaus Elenius, Esa Pitkänen, Relja Popovic, Margarete Fabre, Jennifer Schutzman, Diptee Kulkarni, Alessandro Porello, Andrey Loboda, Heli Lehtonen, Stefan McDonough, Sauli Vuoti, Kai Kaarniranta, Joni A Turunen, Terhi Ollila, Hannu Uusitalo, Juha Karjalainen, Mengzhen Liu, Stephanie Loomis, Erich Strauss, Hao Chen, Kaisa Tasanen, Laura Huilaja, Katariina Hannula-Jouppi, Teea Salmi, Sirkku Peltonen, Leena Koulu, David Choy, Ying Wu, Pirkko Pussinen, Aino Salminen, Tuula Salo, David Rice, Pekka Nieminen, Ulla Palotie, Maria Siponen, Liisa Suominen, Päivi Mäntylä, Ulvi Gursoy, Vuokko Anttonen, Kirsi Sipilä, Rion Pendergrass, Hannele Laivuori, Venla Kurra, Laura Kotaniemi-Talonen, Oskari Heikinheimo, Ilkka Kalliala, Lauri Aaltonen, Varpu Jokimaa, Marja Vääräsmäki, Outi Uimari, Laure Morin-Papunen, Maarit Niinimäki, Terhi Piltonen, Katja Kivinen, Elisabeth Widen, Taru Tukiainen, Niko Välimäki, Eija Laakkonen, Jaakko Tyrmi, Heidi Silven, Eeva Sliz, Riikka Arffman, Susanna Savukoski, Triin Laisk, Natalia Pujol, Janet Kumar, Iiris Hovatta, Erkki Isometsä, Hanna Ollila, Jaana Suvisaari, Antti Mäkitie, Argyro Bizaki-Vallaskangas, Sanna Toppila-Salmi, Tytti Willberg, Elmo Saarentaus, Antti Aarnisalo, Elisa Rahikkala, Kristiina Aittomäki, Fredrik Åberg, Mitja Kurki, Aki Havulinna, Juha Mehtonen, Priit Palta, Shabbeer Hassan, Pietro Della Briotta Parolo, Wei Zhou, Mutaamba Maasha, Susanna Lemmelä, Manuel Rivas, Aoxing Liu, Arto Lehisto, Andrea Ganna, Vincent Llorens, Henrike Heyne, Joel Rämö, Rodos Rodosthenous, Satu Strausz, Tuula Palotie, Kimmo Palin, Javier Garcia-Tabuenca, Harri Siirtola, Tuomo Kiiskinen, Jiwoo Lee, Kristin Tsuo, Kati Hyvärinen, Jarmo Ritari, Katri Pylkäs, Minna Karjalainen, Tuomo Mantere, Eeva Kangasniemi, Sami Heikkinen, Nina Pitkänen, Samuel Lessard, Clément Chatelain, Lila Kallio, Tiina Wahlfors, Eero Punkka, Sanna Siltanen, Teijo Kuopio, Anu Jalanko, Huei-Yi Shen, Risto Kajanne, Mervi Aavikko, Helen Cooper, Denise Öller, Rasko Leinonen, Henna Palin, Malla-Maria Linna, Masahiro Kanai, Zhili Zheng, L. Elisa Lahtela, Mari Kaunisto, Elina Kilpeläinen, Timo P. Sipilä, Oluwaseun Alexander Dada, Awaisa Ghazal, Anastasia Kytölä, Rigbe Weldatsadik, Kati Donner, Anu Loukola, Päivi Laiho, Tuuli Sistonen, Essi Kaiharju, Markku Laukkanen, Elina Järvensivu, Sini Lähteenmäki, Lotta Männikkö, Regis Wong, Auli Toivola, Minna Brunfeldt, Hannele Mattsson, Sami Koskelainen, Tero Hiekkalinna, Teemu Paajanen, Shuang Luo, Shanmukha Sampath Padmanabhuni, Marianna Niemi, Javier Gracia-Tabuenca, Mika Helminen, Tiina Luukkaala, Iida Vähätalo, Jyrki Tammerluoto, Sarah Smith, Tom Southerington, Petri Lehto,

## Notes

### Funding Statement

This study was funded by the following: 
Research Council of Finland (to HM, MH, MT, VL).
Faculty of Health Sciences, University of Eastern Finland (to HM, MT).
Sigrid Juselius Foundation (to MH, VL, ES, TN).
The Strategic Neuroscience Funding of the University of Eastern Finland (to MH, AH, VL, ES).
Alzheimer's Association (to MH).
The State Research Funding KUH-VTR (to VL, ES).
Doctoral Programme in Molecular Medicine (to RMW, HJ).
Deutsche Forschungsgemeinschaft (DFG, German Research Foundation) under Germany's Excellence Strategy within the framework of the Munich Cluster for Systems Nuerology, SyNergy (to CH and SFL) and a Koselleck Project (to CH).
JPco-fuND 2019 Personalised Medicine for Neurodegenerative Diseases; PMG-AD (to CH, SFL, JCL, and MH).

### Author Declarations

Medical Research Ethics committee of Wellbeing Services County of North Savo gave ethical approval for this work.

