## Supplementary material and figures for "Monoallelic *TYROBP* deletion is a novel risk factor for Alzheimer’s disease"

**FinnGen ethics statement**

The FinnGen study is approved by Finnish Institute for Health and Welfare (permit numbers: THL/2031/6.02.00/2017, THL/1101/5.05.00/2017, THL/341/6.02.00/2018, THL/2222/6.02.00/2018, THL/283/6.02.00/2019, THL/1721/5.05.00/2019 and THL/1524/5.05.00/2020), Digital and population data service agency (permit numbers: VRK43431/2017-3, VRK/6909/2018-3, VRK/4415/2019-3), the Social Insurance Institution (permit numbers: KELA 58/522/2017, KELA 131/522/2018, KELA 70/522/2019, KELA 98/522/2019, KELA 134/522/2019, KELA 138/522/2019, KELA 2/522/2020, KELA 16/522/2020), Findata (permit numbers THL/2364/14.02/2020, THL/4055/14.06.00/2020, THL/3433/14.06.00/2020, THL/4432/14.06/2020, THL/5189/14.06/2020, THL/5894/14.06.00/2020, THL/6619/14.06.00/2020, THL/209/14.06.00/2021, THL/688/14.06.00/2021, THL/1284/14.06.00/2021, THL/1965/14.06.00/2021, THL/5546/14.02.00/2020, THL/2658/14.06.00/2021, THL/4235/14.06.00/2021), Statistics Finland (permit numbers: TK-53-1041-17 and TK/143/07.03.00/2020 (earlier TK-53-90-20) TK/1735/07.03.00/2021, TK/3112/07.03.00/2021) and Finnish Registry for Kidney Diseases (permission/extract from the meeting minutes on 4th July 2019).
The Biobank Access Decisions for FinnGen samples and data utilized in FinnGen Data Freeze 12 include: THL Biobank BB2017_55, BB2017_111, BB2018_19, BB_2018_34, BB_2018_67, BB2018_71, BB2019_7, BB2019_8, BB2019_26, BB2020_1, BB2021_65, Finnish Red Cross Blood Service Biobank 7.12.2017, Helsinki Biobank HUS/359/2017, HUS/248/2020, HUS/430/2021 §28, §29, HUS/150/2022 §12, §13, §14, §15, §16, §17, §18, §23, §58, §59, HUS/128/2023 §18, Auria Biobank AB17-5154 and amendment #1 (August 17 2020) and amendments BB_2021-0140, BB_2021-0156 (August 26 2021, Feb 2 2022), BB_2021-0169, BB_2021-0179, BB_2021-0161, AB20-5926 and amendment #1 (April 23 2020) and it´s modifications (Sep 22 2021), BB_2022-0262, BB_2022-0256, Biobank Borealis of Northern Finland_2017_1013, 2021_5010, 2021_5010 Amendment, 2021_5018, 2021_5018 Amendment, 2021_5015, 2021_5015 Amendment, 2021_5015 Amendment_2, 2021_5023, 2021_5023 Amendment, 2021_5023 Amendment_2, 2021_5017, 2021_5017 Amendment, 2022_6001, 2022_6001 Amendment, 2022_6006 Amendment, 2022_6006 Amendment, 2022_6006 Amendment_2, BB22-0067, 2022_0262, 2022_0262 Amendment, Biobank of Eastern Finland 1186/2018 and amendment 22§/2020, 53§/2021, 13§/2022, 14§/2022, 15§/2022, 27§/2022, 28§/2022, 29§/2022, 33§/2022, 35§/2022, 36§/2022, 37§/2022, 39§/2022, 7§/2023, 32§/2023, 33§/2023, 34§/2023, 35§/2023, 36§/2023, 37§/2023, 38§/2023, 39§/2023, 40§/2023, 41§/2023, Finnish Clinical Biobank Tampere MH0004 and amendments (21.02.2020 & 06.10.2020), BB2021-0140 8§/2021, 9§/2021, §9/2022, §10/2022, §12/2022, 13§/2022, §20/2022, §21/2022, §22/2022, §23/2022, 28§/2022, 29§/2022, 30§/2022, 31§/2022, 32§/2022, 38§/2022, 40§/2022, 42§/2022, 1§/2023, Central Finland Biobank 1-2017, BB_2021-0161, BB_2021-0169, BB_2021-0179, BB_2021-0170, BB_2022-0256, BB_2022-0262, BB22-0067, Decision allowing to continue data processing until 31st Aug 2024 for projects: BB_2021-0179, BB22-0067,BB_2022-0262, BB_2021-0170, BB_2021-0164, BB_2021-0161, and BB_2021-0169, and Terveystalo Biobank STB 2018001 and amendment 25th Aug 2020, Finnish Hematological Registry and Clinical Biobank decision 18th June 2021, Arctic biobank P0844: ARC_2021_1001.

Table S1. The 5.2 kb *TYROBP* deletion-associated haplotype identified based on WGS. Variants indicated in bold were included in the final haplotype used to identify putative *TYROBP* deletion carriers in the FinnGen data.

| | | | FinnGen r12 | | gnomAD v4.0.0 | | |
|---|---|---|---|---|---|---|---|
| **rsID** | **Location (GRCh38)** | **Alleles, Ref>Alt** | **MAF** | **INFO** | **MAF Finnish** | **MAF NFE** | **Finnish enrichment** |
| rs12462044 | 19:35837074 | A>G | NA | NA | 0.096 | 0.081 | 1.19 |
| rs12462535 | 19:35837076 | T>A | NA | NA | 0.11 | 0.094 | 1.17 |
| **rs807656** | **19:35847275** | **C>G** | **0.78** | **0.97** | **0.79** | **0.72** | **1.10** |
| **rs1137844** | **19:35852177** | **C>G** | **0.38** | **0.98** | **0.38** | **0.31** | **1.23** |
| **rs11084835** | **19:35886985** | **C>T** | **0.32** | **0.98** | **0.32** | **0.3** | **1.07** |
| **rs1244787406** | **19:35901079** | **T>G** | **0.0022** | **0.84** | **0.0032** | **0.000029** | **110.34** |
| **rs1002399693** | **19:35905673** | **G>C** | **0.0022** | **0.84** | **0.0031** | **0.00003** | **103.33** |

MAF, minor allele frequency; NFE, non-Finnish European.

Table S2. Phenotype associations for rs1244787406 (19:35 901 079 T>G) in FinnGen.

| | OR (95 % CI) | P-value | AF case | AF control |
|---|---|---|---|---|
| **Dementia, including primary healthcare outpatient registry** | 2.10 (1.74-2.52) | 3.29e-15 | 0.0039 | 0.0025 |
| **Dementia** | 2.14 (1.77-2.59) | 4.91e-15 | 0.0039 | 0.0025 |
| **Unspecific neurodegenerative disorder** | 2.20 (1.81-2.68) | 9.75e-15 | 0.0042 | 0.0025 |
| **Any dementia** | 2.20 (1.81-2.68) | 4.76e-14 | 0.0040 | 0.0025 |
| **Alzheimer's disease** | 2.36 (1.87-2.99) | 3.01e-13 | 0.0045 | 0.0025 |

AF, allele frequency.

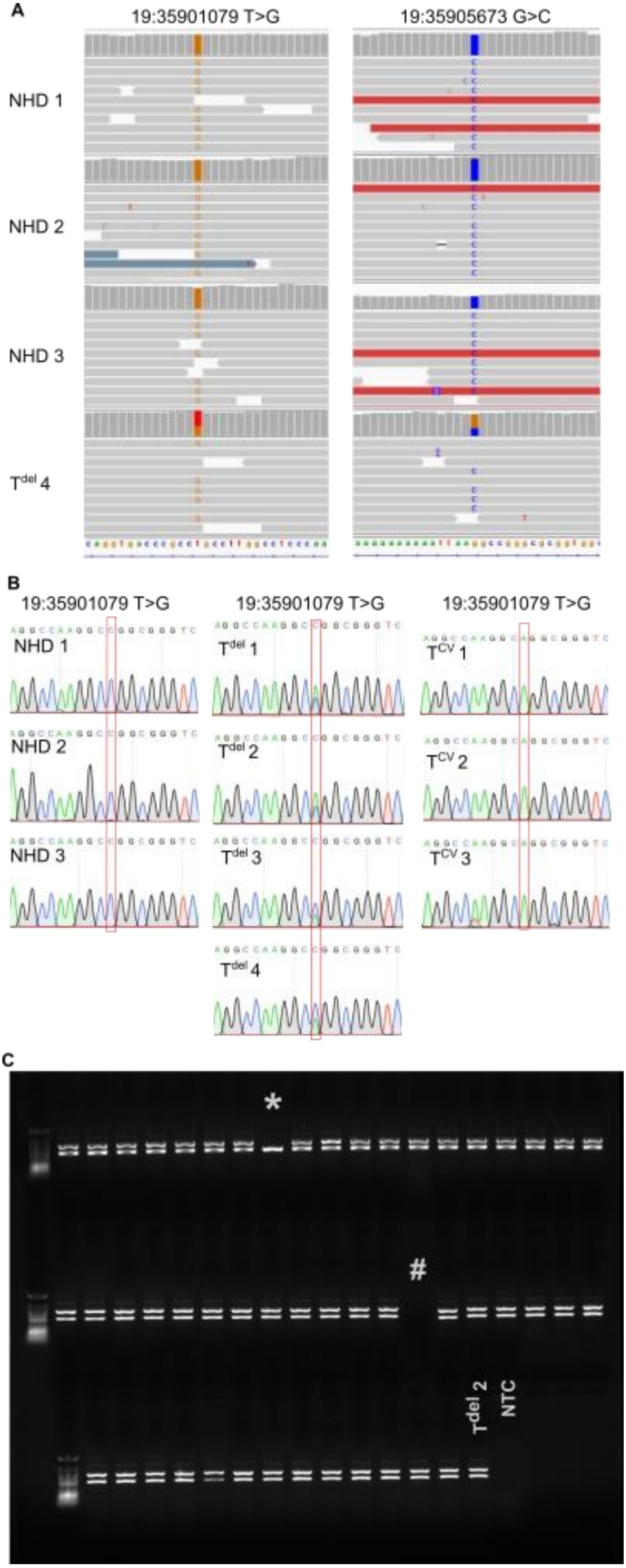


**Fig. S1 Validation of 5.2 kb *TYROBP* deletion proxy markers. (A)** 5.2 kb *TYROBP* deletion proxy markers identified in the whole genome sequencing data of three NHD patients (homozygous for 5.2 kb *TYROBP* deletion) and one monoallelic deletion carrier. **(B)** *TYROBP* deletion proxy marker 19:35901079 T>G was confirmed with Sanger sequencing in three NHD patients, three monoallelic TYROBP deletion carriers and three individuals with common variant of *TYROBP*. **(C)** Deletion specific PCR of 50 imputed 19:35901079 T>G carriers from the FinnGen cohort. Two bands (~500 kb and ~700 kb) indicate individuals heterozygous for the *TYROBP* deletion, while single 500 kb band indicates individual homozygous for the common variant of *TYROBP*. * denotes an individual with low (0.52) genotype probability for 19:35901079 T>G variant. # denotes an empty well. NHD, Nasu Hakola disease patient; NTC, non-template control; $T^{del}$, monoallelic *TYROBP* deletion carrier; $T^{CV}$, *TYROBP* common variant.

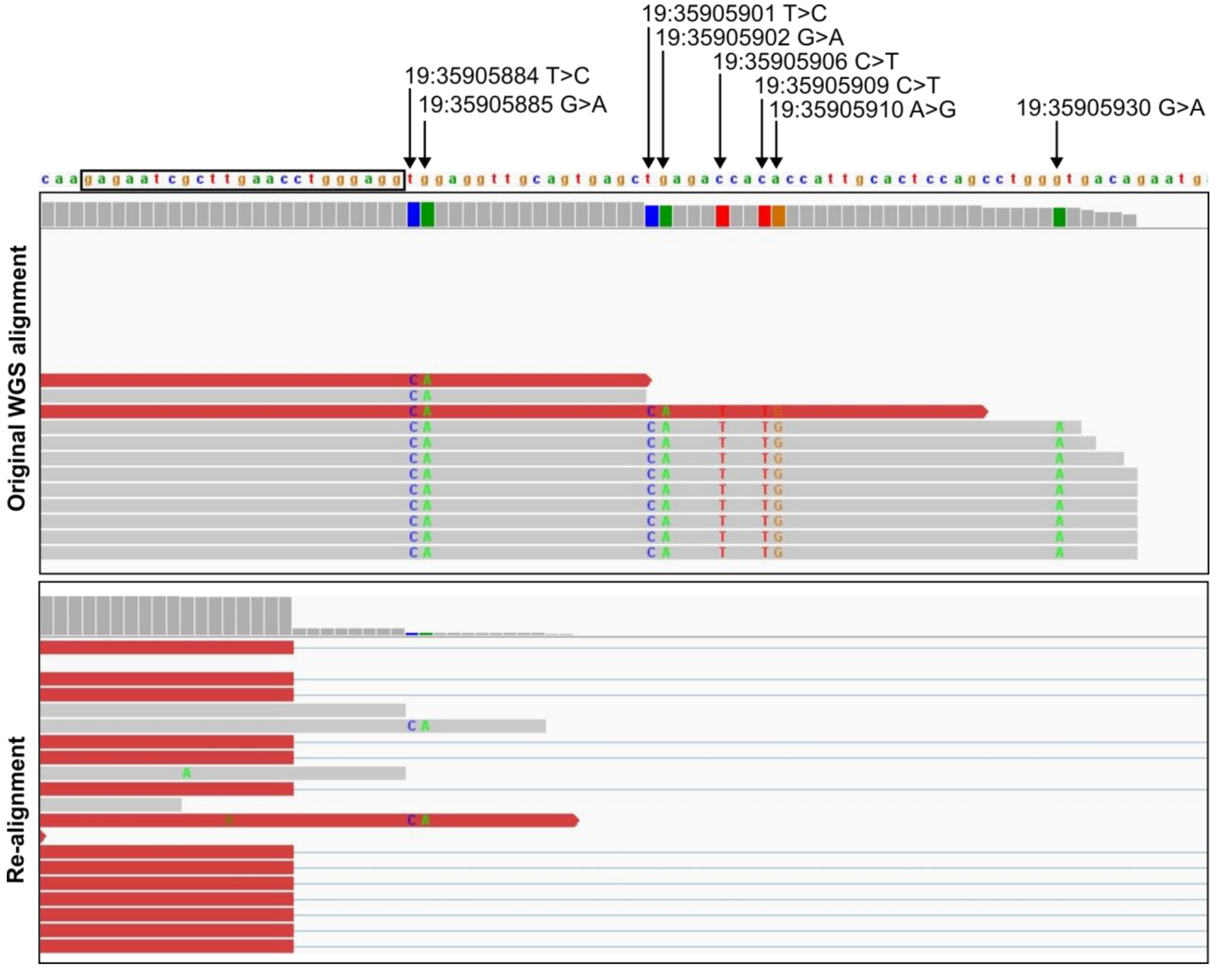


**Fig. S2 Artefact single nucleotide variants arise from incorrect alignment of WGS reads spanning the 5.2 kb *TYROBP* deletion.** The upper panel shows the original alignment of WGS data from an NHD patient with homozygous 5.2 kb *TYROBP* deletion. The lower panel shows the same data after re-alignment with parameters allowing large deletions. Horizontal lines in the lower panel indicate reads that span across the deletion. The deletion 5’ and 3’ break points are located within a 23 bp identical sequence (indicated in the figure with a black box) within 120 bp almost identical *Alu* repeats. This sequence similarity leads to incorrect alignment of the reads that span the deletion when standard parameters for WGS read alignment are used and leads to detection of artefact SNVs at chromosome positions within the deleted region (indicated by arrows and chromosomal location in the figure).

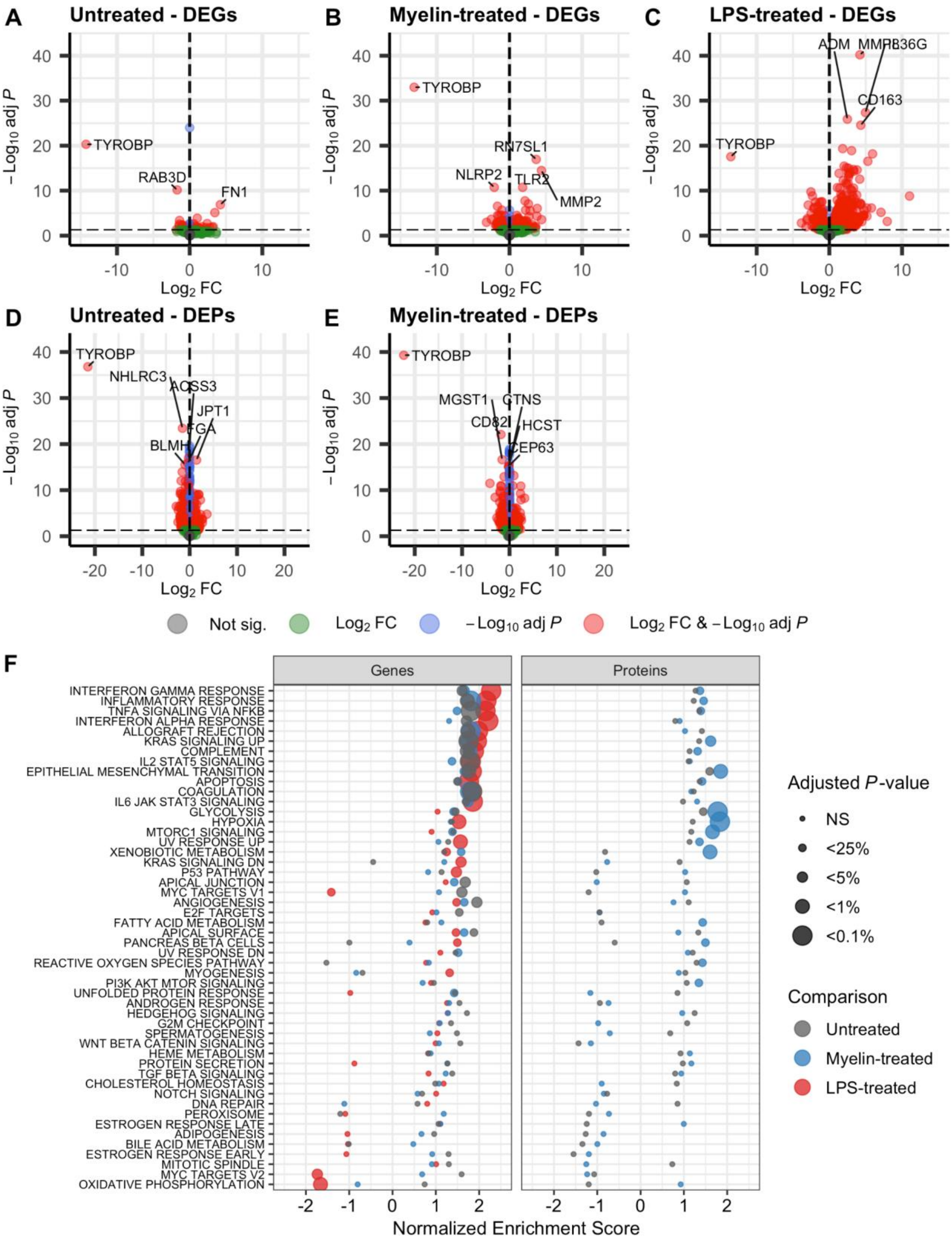


**Fig S3 Increased inflammatory response induced by biallelic *TYROBP* deletion in MDMi cells. (A-C)** Differentially expressed genes (DEG) and **(D-E)** proteins (DEP) in NHD patient MDMi cells compared to controls upon untreated (A, D), myelin-treated (B, E) and LPS-treated (C) conditions. **(F)** Pathway enrichment of genes (left panel) and proteins (right panel) differentially expressed in NHD patient MDMi cells compared to controls. NHD, n=2 (A-E); controls with *TYROBP* common variant, n=12 (A), n=7 (B), n=3 (C-E).

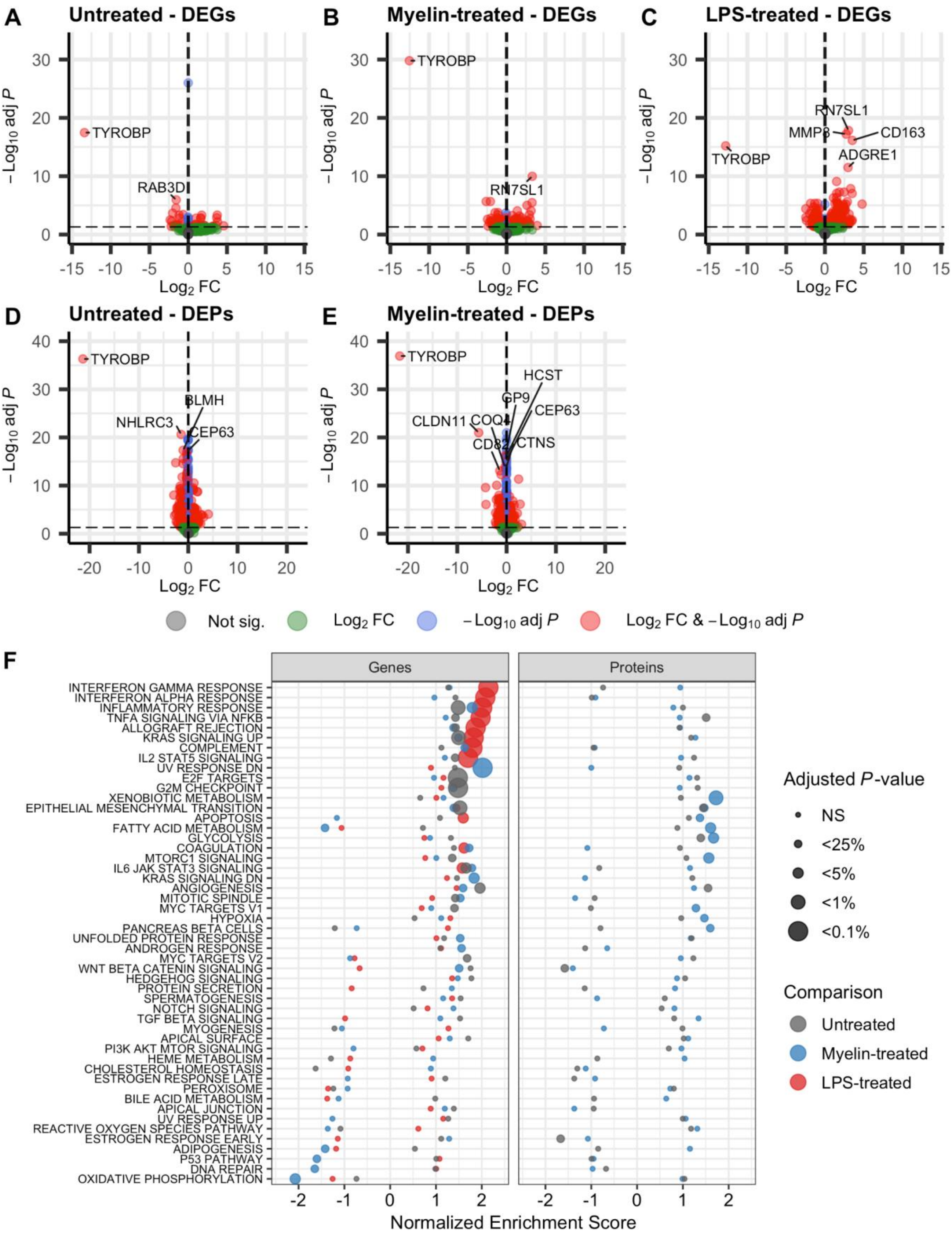


**Fig S4 Altered pathways induced by biallelic vs. monoallelic *TYROBP* deletion in MDMi cells. (A-C)** Differentially expressed genes (DEG) and **(D-E)** proteins (DEP) in NHD patient MDMi cells compared to monoallelic *TYROBP* deletion carriers upon untreated (A, D), myelin-treated (B, E) and LPS-treated (C) conditions. **(F)** Pathway enrichment of genes (left panel) and proteins (right panel) differentially expressed in NHD patient MDMi cells compared to monoallelic *TYROBP* deletion carriers. NHD, n=2 (A-E); monoallelic *TYROBP* deletion carrier, n=3 (A-E).

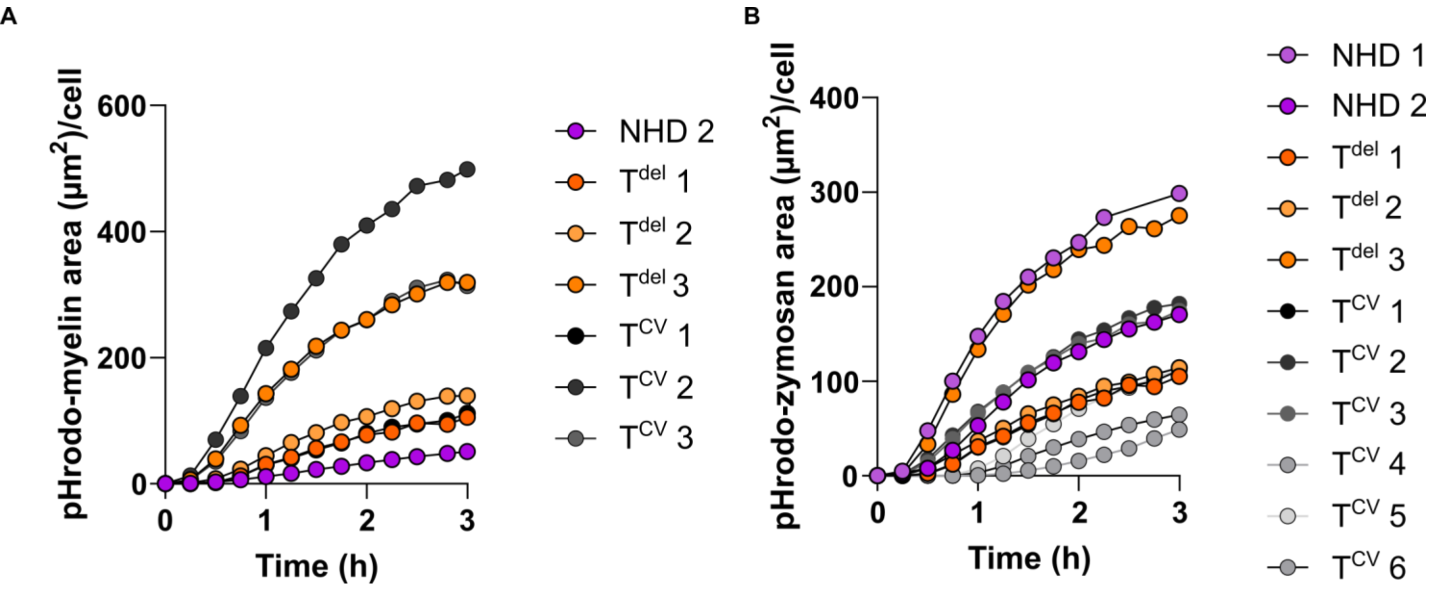


**Fig S5 (A-B)** Phagocytic activity of $T^{CV}$, $T^{del}$, and NHD MDMi was assessed by challenging the cells with pHrodo-labelled myelin or zymosan. Images were obtained every 15 min for a total of 3 hours. pHrodo positive area was quantified and normalized to cell count. Each data point represents an average of 1-3 replicate wells.

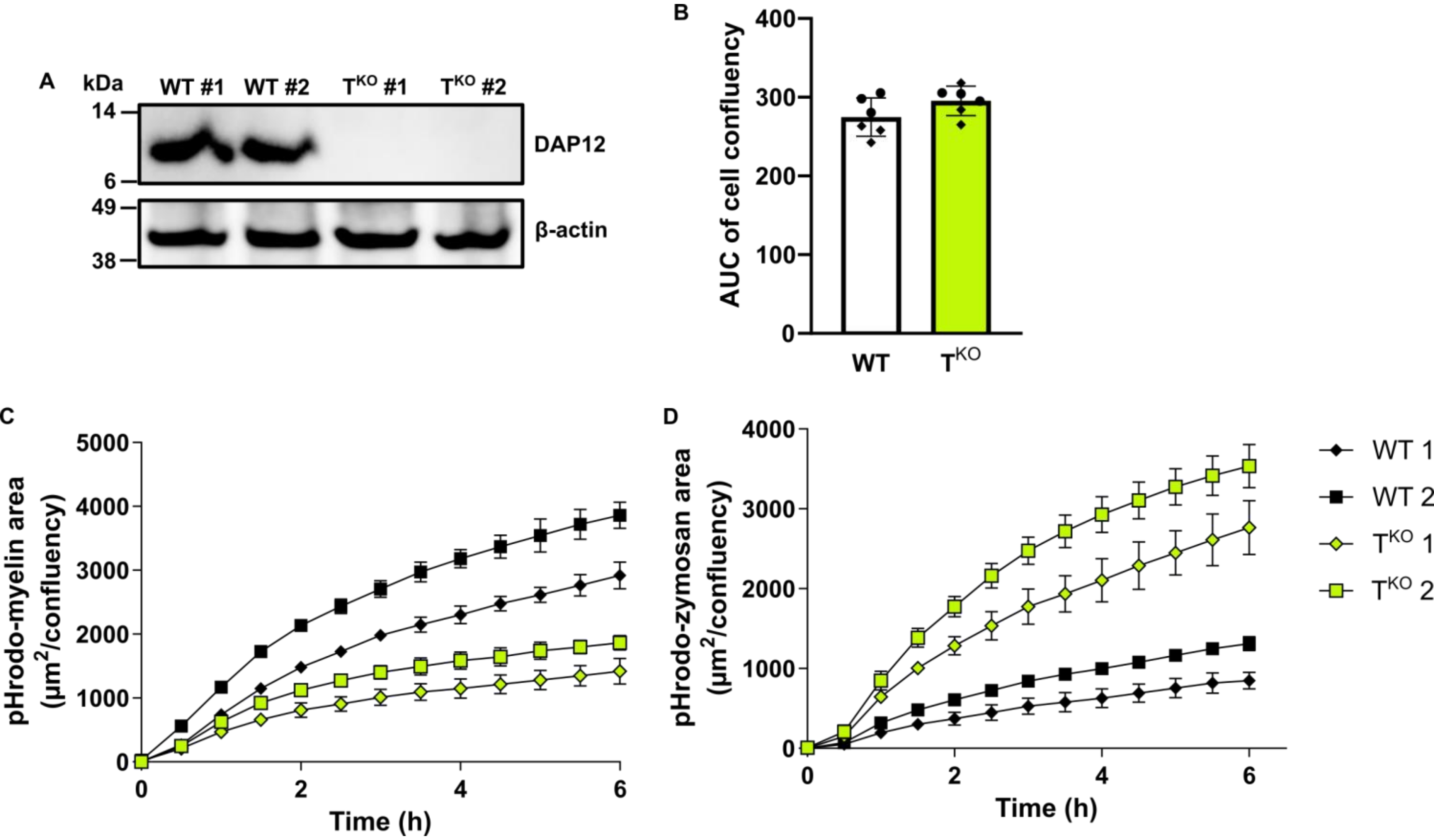


**Fig S6 Validation of *Tyrobp* KO BV2 cell lines. (A)** Western blot of cell lysates from two *Tyrobp* KO lines and two wild type control lines confirms loss of DAP12 protein in the KO lines, while DAP12 is robustly expressed by the wild type lines. **(B)** Cell growth was monitored for 24 h by live cell imaging. Cell confluency at different time points was measured, and an area under the curve (AUC) was calculated. No difference in the AUC of cell confluency between wild type and *Tyrobp* KO lines was observed (independent samples t-test). **(C-D)** Phagocytic activity in BV2 cells was assessed by quantifying the area of pHrodo-labelled myelin and zymosan signal normalized to confluency. Different symbols represent the different lines, each symbol represents one technical replicate (three technical replicates from each line).
